# Early potassium response to mineralocorticoid receptor antagonist therapy and cardiovascular outcomes in primary aldosteronism: a landmark cohort study

**DOI:** 10.64898/2026.11.15.26368152

**Authors:** Szu-Han Chen, Yen Hsun Huang, Yu-Hsuan Lee, Shu-Mei Yang, Chi-Jung Huang, Yuan-Kai Pang, Yun-Yu Chen, Hao-Min Cheng

## Abstract

**Background:** Mineralocorticoid receptor antagonists (MRAs) are standard therapy for primary aldosteronism (PA). Serum potassium is measured in nearly every treated patient, but the long-term prognostic value of potassium attained early after initiation is undefined.

**Methods:** In the TriNetX Research Network, we identified adults with PA newly initiating spironolactone or eplerenone in 2000–2025, without prior adrenalectomy (n=4,169; 180-day landmark design). Achieved potassium (last value on days 14–180) was categorised as <4.0, 4.0–<4.5, 4.5–<5.0 and ≥5.0 mmol/L, and its change from baseline in tertiles, using cause-specific Cox and Fine-Gray models. The primary outcome was a composite of myocardial infarction, stroke or death; incident heart failure was co-primary.

**Findings:** Over a median 3.0 years, 623 composite events occurred. Estimated risk was lowest at 4.5–<5.0 mmol/L (adjusted hazard ratio 0.65, 95% CI 0.52–0.83, versus <4.0 mmol/L), lower than the 4.0–<4.5 mmol/L band, and not different from ≥5.0 mmol/L. A larger rise in potassium was associated with lower risk. A similar pattern was seen for incident heart failure (0.54, 0.36–0.80; 248 events). Potassium below the 3.6 mmol/L consensus threshold for hypokalaemia carried no significantly higher risk (1.22, 0.96–1.56). The change in potassium correlated weakly with the change in renin (ρ=0.15).

**Interpretation:** In MRA-treated PA, cardiovascular and heart-failure risk was lowest at an early achieved potassium of 4.5–<5.0 mmol/L, well above the 3.6 mmol/L accepted by consensus criteria as correction of hypokalaemia; correction alone may therefore be an insufficient treatment target. Whether titrating therapy towards this band improves outcomes requires prospective evaluation.

**Funding:** Grants from Taipei Veterans General Hospital (V114C-038) and the National Science and Technology Council, Taiwan (NSTC 113-2314-B-A49-040-MY3, 113-2314-B-A49-009, 113-2321-B-075A-002).

## Introduction

Primary aldosteronism (PA) is the most common endocrine cause of hypertension, and the cardiovascular risk it carries is not explained by blood pressure (BP) alone. Compared with essential hypertension at similar BP, it is associated with higher risks of stroke (odds ratio 2.58, 95% CI 1.93–3.45), coronary artery disease, atrial fibrillation, and heart failure (HF; odds ratio 2.05, 95% CI 1.11–3.78),^1^ and that excess is not abolished by treatment: among patients treated with mineralocorticoid receptor antagonists (MRAs), the adjusted hazard of incident cardiovascular events remains almost twice that of matched patients with essential hypertension.^2^ Hypokalaemia, the feature that most often prompts testing, is present in only a minority of cases,^3^ so potassium serves less as a diagnostic marker than as the parameter monitored in nearly every treated patient.

What constitutes an adequate response to medical treatment therefore has direct prognostic consequences, and it remains unsettled. The Primary Aldosteronism Medical Treatment Outcome (PAMO) consensus defines a complete biochemical response as correction of hypokalaemia without potassium supplementation together with normalisation of renin, and found that only about half of medically treated patients achieve it.^4^ Its potassium criterion, like the rest of the definition, was set by Delphi consensus: correction is defined as a serum potassium of at least 3.6 mmol/L, the same threshold used for biochemical success after adrenalectomy in the Primary Aldosteronism Surgical Outcome (PASO) consensus,^5^ and one that has not, to our knowledge, been validated against cardiovascular outcomes. Renin is the better-evidenced component: the excess risk is concentrated in patients whose renin remains suppressed,^2^ unsuppressed post-treatment renin was associated with fewer cardiovascular events (pooled hazard ratio [HR] 0.43, 95% confidence interval [CI] 0.23–0.80),^6^ and the 2025 Endocrine Society guideline recommends monitoring renin and titrating the MRA to raise it when suppressed.^7^ Renin, however, is measured in a minority of patients in routine care. Mineralocorticoid receptor activation drives myocardial fibrosis and adverse remodelling,^8^ and blockade reduces HF in trials outside PA,^9,10^ so HF is the outcome in which an adequate blockade signal might be most visible.

Potassium occupies an unusual position in this framework. A 2026 review ranks its normalisation second among the three objectives of medical therapy, while stating that no specific target level exists for PA.^11^ The target levels that exist come from other populations, where the potassium–mortality relationship is U-shaped and lowest at about 4.0–4.5 mmol/L^11–15^ — so a higher achieved concentration cannot be assumed to be better. We therefore used a landmark design to examine whether the early potassium response to MRA therapy, expressed both as the achieved concentration and as the change from baseline, is associated with subsequent cardiovascular events and incident HF in adults with PA, whether such association is confined to the correction of frank hypokalaemia, and how it relates to the change in renin.

## Methods

### Data Source

Data were obtained from the TriNetX Research Network, a federated electronic health record network providing de-identified clinical data from 91 contributing healthcare organisations, yielding 10,661 unique patients (10,077 after the diagnosis-code exclusions below). Individual informed consent was waived under Health Insurance Portability and Accountability Act (HIPAA). The study was approved by the Institutional Review Board of Taichung Veterans General Hospital (CE251118C) and follows the RECORD extension of STROBE (Supplementary Table S1).

### Study Population

We identified adults with PA newly initiating spironolactone or eplerenone, without a prior MRA prescription or adrenalectomy. PA was ascertained by ICD-10-CM codes beginning E26 (hyperaldosteronism), excluding E26.1 (secondary hyperaldosteronism) and E26.81 (Bartter syndrome), neither of which is PA (code list in Supplementary Table S2); the unspecified codes E26 and E26.9 were retained. The index date was the first prescription of either agent. The case definition remains administrative, with no requirement for confirmatory testing: about half the analytic cohort carried a code specific to PA and most of the remainder only unspecified hyperaldosteronism.

### Design and Exposure

We used a landmark design to separate measurement of the early biochemical response from the accrual of outcomes. Baseline potassium was the last value in the 365 days before the index date; achieved potassium was the last value on days 14 to 180, and the landmark was day 180. Follow-up was measured from the landmark, and events before it were exclusions rather than outcomes. Achieved potassium was categorised as <4.0, 4.0 to <4.5, 4.5 to <5.0 and ≥5.0 mmol/L, with <4.0 as the reference; the change from baseline (ΔK) was analysed in tertiles. Patients were required to have paired potassium values, to be free of end-stage renal disease and of a primary-outcome event at the landmark, to remain under observation, and to have complete covariate data (Figure 1; Supplementary Table S3). Details of measurement timing, specimen types and selection weighting are in the Supplementary Methods.

**Figure 1.**
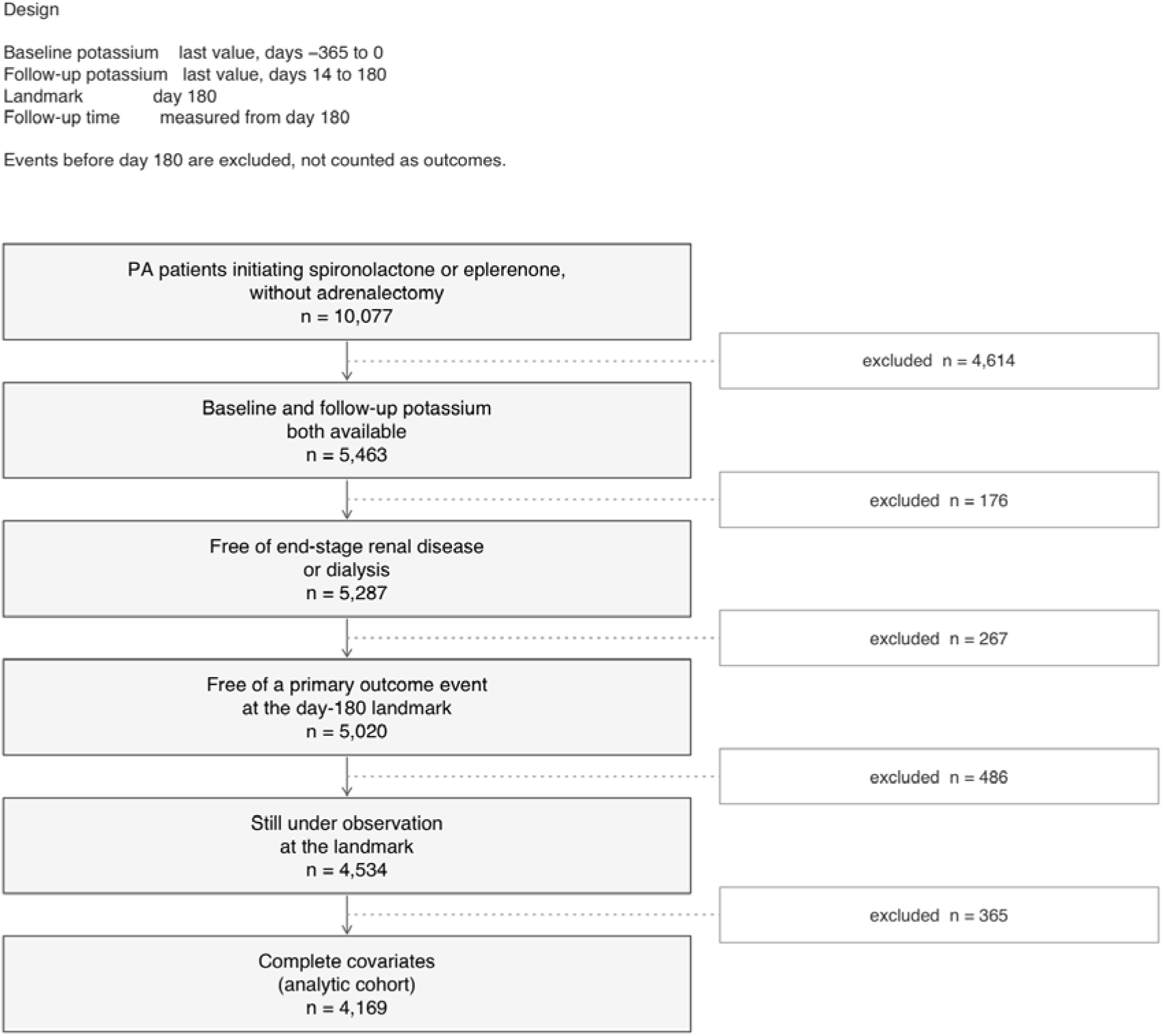
Cohort derivation. Baseline potassium: last value, days −365 to 0. Follow-up potassium: last value, days 14 to 180. Landmark: day 180. Events before day 180 are excluded, not counted as outcomes; follow-up time is measured from day 180.

**Figure 2.**
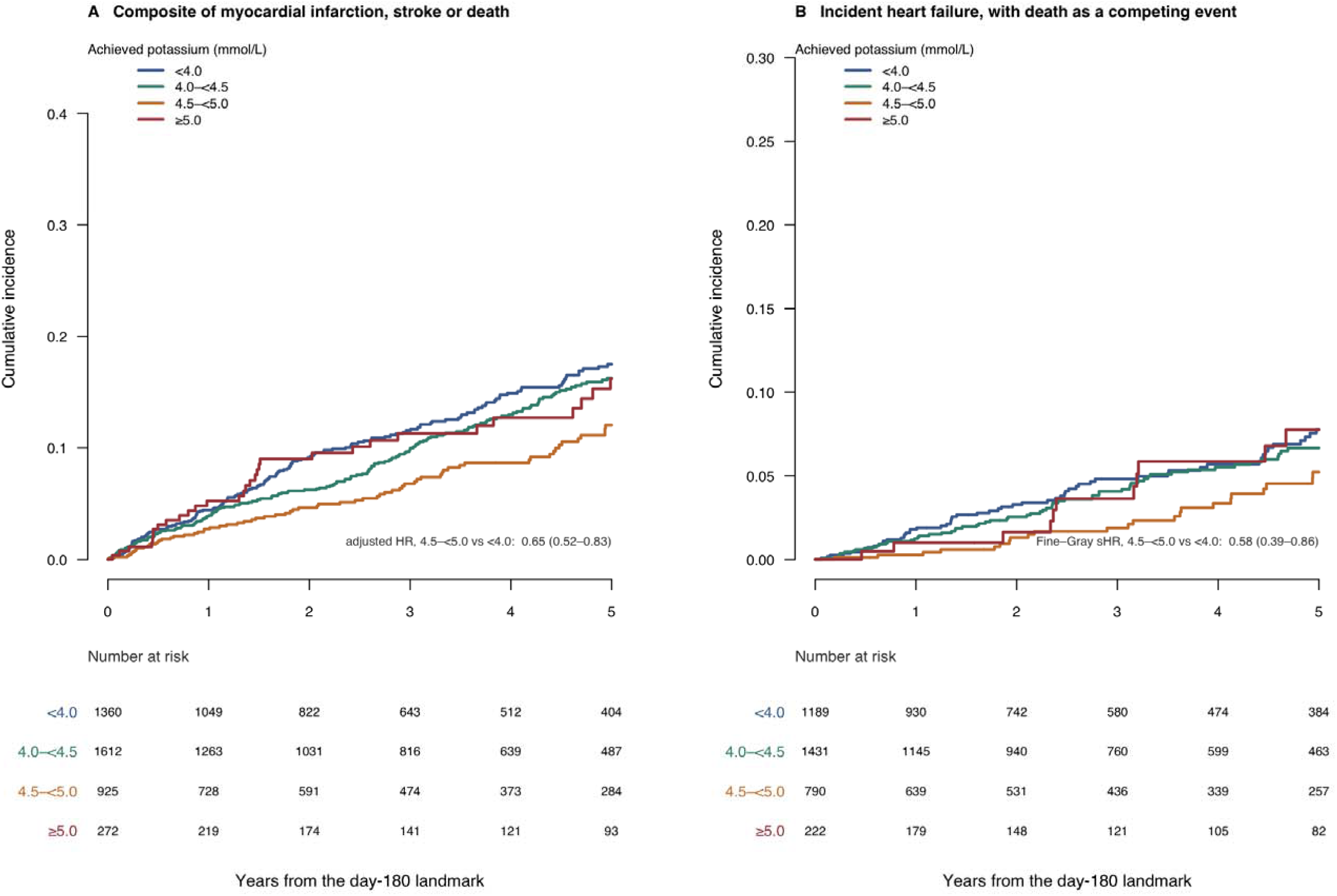
Cumulative incidence of (A) the composite outcome and (B) incident heart failure, by achieved potassium category, from the day-180 landmark. Curves are unadjusted: panel A shows 1 minus the Kaplan–Meier estimate, and panel B the Aalen–Johansen cumulative incidence with death as a competing event. The estimate printed in each panel comes from a separate adjusted model: a cause-specific hazard ratio (HR) in panel A and a Fine–Gray subdistribution hazard ratio (sHR) in panel B, each for 4.5–<5.0 versus <4.0 mmol/L. Numbers at risk are shown below each panel.

### Outcomes

The primary outcome was a composite of myocardial infarction, stroke and all-cause death. Incident HF was a co-primary outcome, analysed in the subset free of HF both at baseline and through the landmark. Non-fatal outcomes were required to be coded at an inpatient, acute-care or emergency encounter, and follow-up continued until the outcome, death, or the last recorded encounter. Because all-cause death contributes a substantial share of composite events and its ascertainment varies by calendar era in this network, we additionally report the composite restricted to myocardial infarction and stroke, and examine the association by calendar period of MRA initiation.

### Covariates

Models were adjusted for age, sex, race, MRA agent, baseline potassium, baseline estimated glomerular filtration rate (eGFR), hypertension, diabetes, chronic kidney disease (CKD), atrial fibrillation and ischemic heart disease, and for concomitant medications recorded from 180 days before the index date through the landmark: angiotensin-converting-enzyme inhibitors, angiotensin receptor blockers, thiazides, loop diuretics and potassium supplements. Models were stratified on prevalent cerebrovascular disease (ICD-10 I60–I69) and, for the composite outcome, on prevalent HF. eGFR was estimated with the 2021 CKD-EPI creatinine equation. Race was categorised as recorded in the electronic health record (White, Black or African American, Asian, other or unknown) and was retained as a covariate because it was associated with both achieved potassium and outcome; models with and without race are compared in Supplementary Table S31.

ΔeGFR and achieved BP are measured in the exposure window and were therefore excluded from the primary models and examined in sensitivity analyses (S11, S34); all estimates are multivariable-adjusted associations rather than causal effects.

### Statistical Analysis

The proportional hazards assumption was assessed via scaled Schoenfeld residuals for every model term and an explicit interaction between exposure and a split of follow-up at three years. Fine-Gray subdistribution models, treating death as a competing event, were fitted as a sensitivity analysis for incident HF. Three-year cumulative incidence from the landmark was estimated unadjusted by the Aalen-Johansen estimator and, adjusted, by g-formula standardisation over the fitted model, with percentile confidence intervals from 500 nonparametric bootstrap replicates.

Departure from a linear dose-response was examined by nested likelihood-ratio tests comparing a linear term against fractional polynomials and restricted cubic splines, with and without a knot at 5.0 mmol/L; these are global tests of fit, not tests of where a departure lies. Because the reference category pools frank hypokalaemia with low-normal potassium, we also split it at 3.5 mmol/L, repeated the analysis restricted to the reference range (3.5 to <5.0 mmol/L), and estimated the contrast between adjacent categories. Because the PAMO consensus defines correction of hypokalaemia as a serum potassium of at least 3.6 mmol/L, achieved potassium was also dichotomised at 3.6 mmol/L and categorised with 3.6 to <4.0 mmol/L as the reference; among patients with a baseline potassium below 3.6 mmol/L, the three PAMO potassium responses (corrected without supplementation, corrected with supplementation, not corrected) were compared, with supplementation defined as a potassium prescription between the index date and the landmark.

Because renin is the established marker of response to mineralocorticoid blockade, we assembled a bridging sample in which plasma renin activity was available both before MRA initiation and in the days 14–180 window, in the same assay units and paired with potassium from the same windows. Within it we estimated the Spearman correlation between the change in potassium and the change in renin, and the discrimination of an unsuppressed renin at follow-up by the area under the receiver operating characteristic curve. Renin was classified as unsuppressed at a plasma renin activity of 1.0 ng/mL/h or greater;^6^ because a suppressed pre-treatment value was not required for entry, this describes renin status at follow-up rather than a transition from suppressed to unsuppressed.

Robustness was examined in pre-specified subgroups and 12 analytic variations listed in the Supplementary Methods, including a stricter two-diagnosis-code requirement (evaluated because two separately dated codes raise reproducibility without necessarily raising specificity). Cataract, herpes zoster and inguinal hernia served as negative control outcomes; achieved systolic BP served as a positive control. E-values were calculated with the common-outcome correction.

Two-sided P values are reported; because many secondary and sensitivity analyses were performed, these are interpreted as supportive rather than confirmatory, and no adjustment for multiplicity was made except where stated. Analyses were performed in R version 4.6.1 (R Foundation for Statistical Computing, Vienna, Austria) using the survival (version 3.8.6) package. The fractional-polynomial search and E-values used study-specific code rather than a dedicated package; bootstrap standardisation used the boot package. Analysis code is available from the corresponding author on reasonable request.

## Results

### Cohort and Early Potassium Response

Of 10,077 adults with PA newly initiating spironolactone or eplerenone, 4,169 had paired potassium measurements, were free of end-stage renal disease, remained event-free and under observation at the day-180 landmark, and had complete covariate data (Figure 1). Over a median follow-up of 3.0 years from the landmark, 623 composite events of myocardial infarction, stroke, or death occurred; 183 patients had a myocardial infarction, 183 a stroke and 379 died, counts that overlap because only the first event contributes.

Baseline and early post-initiation characteristics are shown in Table 1. Median baseline potassium was 3.8 mmol/L (IQR, 3.4–4.1) and median achieved potassium 4.2 mmol/L (IQR, 3.8–4.5), a median rise of 0.3 mmol/L (IQR, 0.0–0.8). Patients reaching higher achieved potassium were progressively older, had lower baseline eGFR, more often had CKD, were less likely to be Black, and were less often taking a potassium supplement.

**Table 1.**
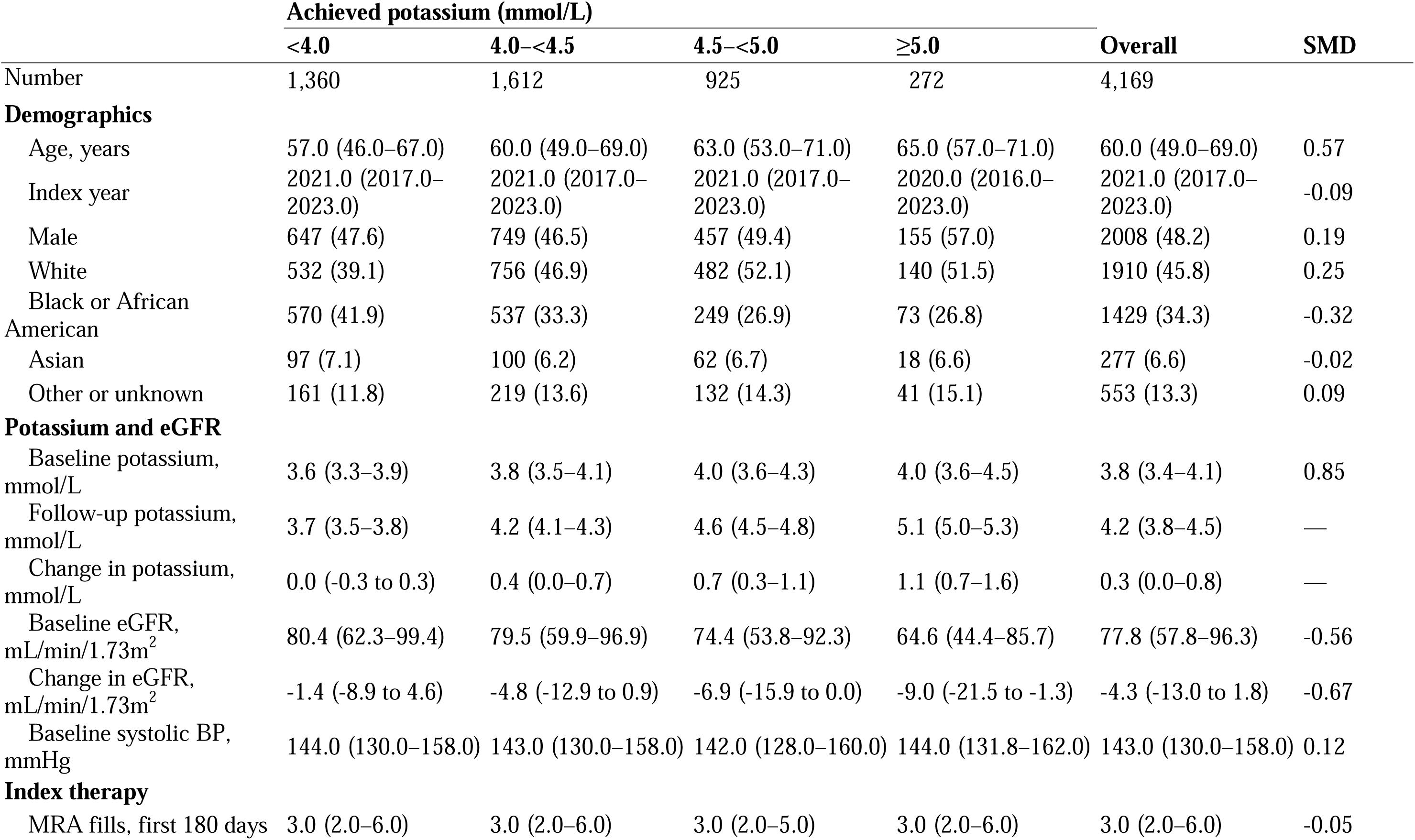
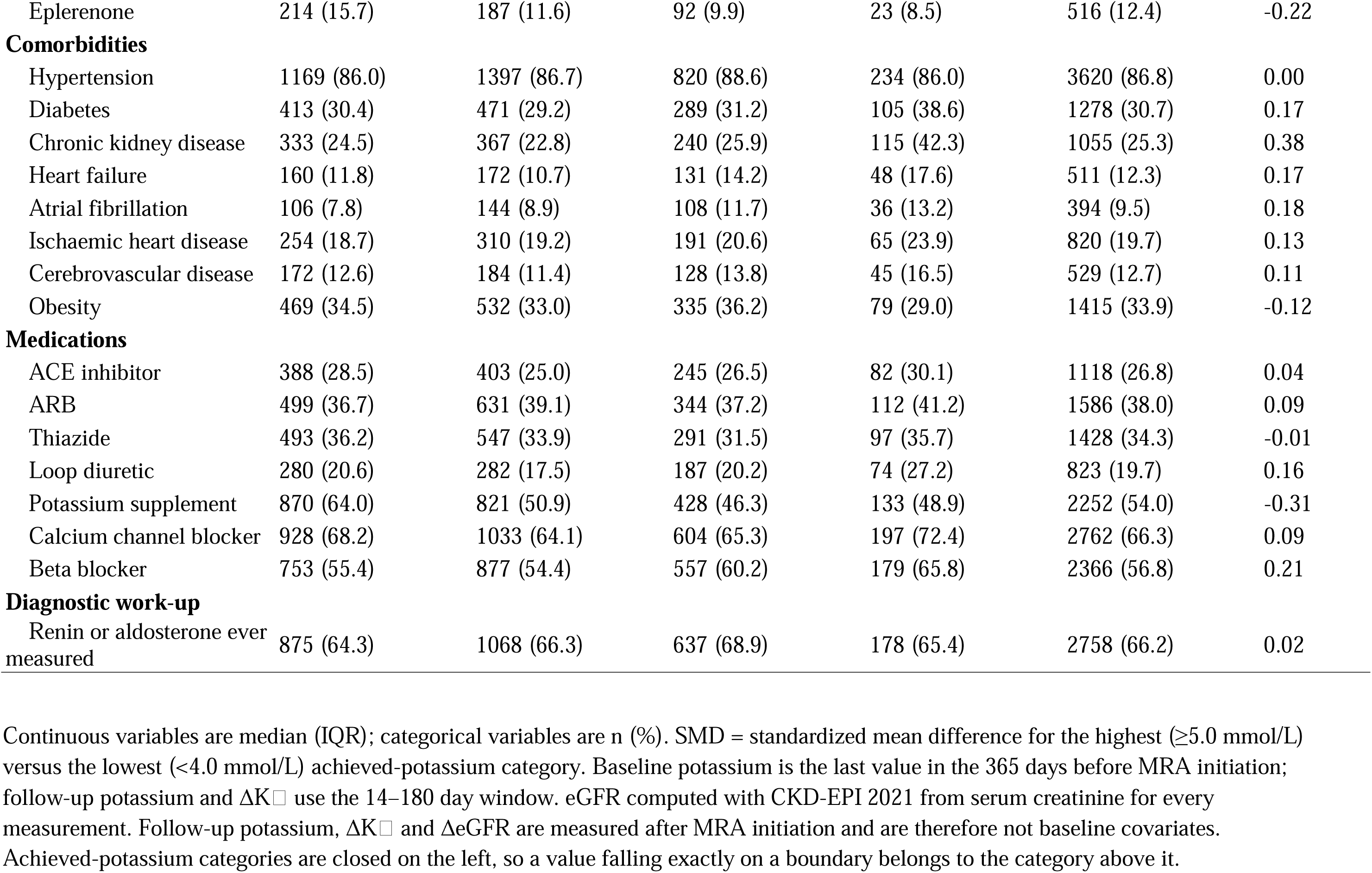
Baseline and early post-initiation characteristics of the analytic cohort (n = 4,169), by achieved potassium category 14–180 days after MRA initiation.

|  | **Achieved potassium (mmol/L)** |  |  |  |  |  |
|---|---|---|---|---|---|---|
|  | **<4.0** | **4.0–<4.5** | **4.5–<5.0** | **≥5.0** | **Overall** | **SMD** |
| Number | 1,360 | 1,612 | 925 | 272 | 4,169 |  |
| **Demographics** |  |  |  |  |  |  |
| Age, years | 57.0 (46.0–67.0) | 60.0 (49.0–69.0) | 63.0 (53.0–71.0) | 65.0 (57.0–71.0) | 60.0 (49.0–69.0) | 0.57 |
| Index year | 2021.0 (2017.0–2023.0) | 2021.0 (2017.0–2023.0) | 2021.0 (2017.0–2023.0) | 2020.0 (2016.0–2023.0) | 2021.0 (2017.0–2023.0) | -0.09 |
| Male | 647 (47.6) | 749 (46.5) | 457 (49.4) | 155 (57.0) | 2008 (48.2) | 0.19 |
| White | 532 (39.1) | 756 (46.9) | 482 (52.1) | 140 (51.5) | 1910 (45.8) | 0.25 |
| Black or African American | 570 (41.9) | 537 (33.3) | 249 (26.9) | 73 (26.8) | 1429 (34.3) | -0.32 |
| Asian | 97 (7.1) | 100 (6.2) | 62 (6.7) | 18 (6.6) | 277 (6.6) | -0.02 |
| Other or unknown | 161 (11.8) | 219 (13.6) | 132 (14.3) | 41 (15.1) | 553 (13.3) | 0.09 |
| **Potassium and eGFR** |  |  |  |  |  |  |
| Baseline potassium, mmol/L | 3.6 (3.3–3.9) | 3.8 (3.5–4.1) | 4.0 (3.6–4.3) | 4.0 (3.6–4.5) | 3.8 (3.4–4.1) | 0.85 |
| Follow-up potassium, mmol/L | 3.7 (3.5–3.8) | 4.2 (4.1–4.3) | 4.6 (4.5–4.8) | 5.1 (5.0–5.3) | 4.2 (3.8–4.5) | — |
| Change in potassium, mmol/L | 0.0 (-0.3 to 0.3) | 0.4 (0.0–0.7) | 0.7 (0.3–1.1) | 1.1 (0.7–1.6) | 0.3 (0.0–0.8) | — |
| Baseline eGFR, mL/min/1.73m<sup>2</sup> | 80.4 (62.3–99.4) | 79.5 (59.9–96.9) | 74.4 (53.8–92.3) | 64.6 (44.4–85.7) | 77.8 (57.8–96.3) | -0.56 |
| Change in eGFR, mL/min/1.73m<sup>2</sup> | -1.4 (-8.9 to 4.6) | -4.8 (-12.9 to 0.9) | -6.9 (-15.9 to 0.0) | -9.0 (-21.5 to -1.3) | -4.3 (-13.0 to 1.8) | -0.67 |
| Baseline systolic BP, mmHg | 144.0 (130.0–158.0) | 143.0 (130.0–158.0) | 142.0 (128.0–160.0) | 144.0 (131.8–162.0) | 143.0 (130.0–158.0) | 0.12 |
| **Index therapy** |  |  |  |  |  |  |
| MRA fills, first 180 days | 3.0 (2.0–6.0) | 3.0 (2.0–6.0) | 3.0 (2.0–5.0) | 3.0 (2.0–6.0) | 3.0 (2.0–6.0) | -0.05 |
| Eplerenone | 214 (15.7) | 187 (11.6) | 92 (9.9) | 23 (8.5) | 516 (12.4) | -0.22 |
| **Comorbidities** |  |  |  |  |  |  |
| Hypertension | 1169 (86.0) | 1397 (86.7) | 820 (88.6) | 234 (86.0) | 3620 (86.8) | 0.00 |
| Diabetes | 413 (30.4) | 471 (29.2) | 289 (31.2) | 105 (38.6) | 1278 (30.7) | 0.17 |
| Chronic kidney disease | 333 (24.5) | 367 (22.8) | 240 (25.9) | 115 (42.3) | 1055 (25.3) | 0.38 |
| Heart failure | 160 (11.8) | 172 (10.7) | 131 (14.2) | 48 (17.6) | 511 (12.3) | 0.17 |
| Atrial fibrillation | 106 (7.8) | 144 (8.9) | 108 (11.7) | 36 (13.2) | 394 (9.5) | 0.18 |
| Ischaemic heart disease | 254 (18.7) | 310 (19.2) | 191 (20.6) | 65 (23.9) | 820 (19.7) | 0.13 |
| Cerebrovascular disease | 172 (12.6) | 184 (11.4) | 128 (13.8) | 45 (16.5) | 529 (12.7) | 0.11 |
| Obesity | 469 (34.5) | 532 (33.0) | 335 (36.2) | 79 (29.0) | 1415 (33.9) | -0.12 |
| **Medications** |  |  |  |  |  |  |
| ACE inhibitor | 388 (28.5) | 403 (25.0) | 245 (26.5) | 82 (30.1) | 1118 (26.8) | 0.04 |
| ARB | 499 (36.7) | 631 (39.1) | 344 (37.2) | 112 (41.2) | 1586 (38.0) | 0.09 |
| Thiazide | 493 (36.2) | 547 (33.9) | 291 (31.5) | 97 (35.7) | 1428 (34.3) | -0.01 |
| Loop diuretic | 280 (20.6) | 282 (17.5) | 187 (20.2) | 74 (27.2) | 823 (19.7) | 0.16 |
| Potassium supplement | 870 (64.0) | 821 (50.9) | 428 (46.3) | 133 (48.9) | 2252 (54.0) | -0.31 |
| Calcium channel blocker | 928 (68.2) | 1033 (64.1) | 604 (65.3) | 197 (72.4) | 2762 (66.3) | 0.09 |
| Beta blocker | 753 (55.4) | 877 (54.4) | 557 (60.2) | 179 (65.8) | 2366 (56.8) | 0.21 |
| **Diagnostic work-up** |  |  |  |  |  |  |
| Renin or aldosterone ever measured | 875 (64.3) | 1068 (66.3) | 637 (68.9) | 178 (65.4) | 2758 (66.2) | 0.02 |

Continuous variables are median (IQR); categorical variables are n (%). SMD = standardized mean difference for the highest (≥5.0 mmol/L) versus the lowest (<4.0 mmol/L) achieved-potassium category. Baseline potassium is the last value in the 365 days before MRA initiation; follow-up potassium and ΔK□ use the 14–180 day window. eGFR computed with CKD-EPI 2021 from serum creatinine for every measurement. Follow-up potassium, ΔK□ and ΔeGFR are measured after MRA initiation and are therefore not baseline covariates. Achieved-potassium categories are closed on the left, so a value falling exactly on a boundary belongs to the category above it.

### Achieved Potassium and the Composite Outcome

Estimated risk was lowest at an achieved potassium of 4.5–<5.0 mmol/L, with higher point estimates on either side of it (Table 2). Relative to <4.0 mmol/L, the adjusted HR was 0.65 (95% CI, 0.52–0.83; P < 0.001) for 4.5–<5.0 mmol/L. The two remaining categories did not differ significantly from the reference: 0.88 (95% CI, 0.73–1.07; P = 0.191) for 4.0–<4.5 mmol/L and 0.80 (95% CI, 0.59–1.09; P = 0.159) for ≥5.0 mmol/L, the latter resting on 272 patients with 58 events. With 4.0–<4.5 mmol/L as the reference, 4.5–<5.0 mmol/L gave 0.74 (95% CI, 0.59–0.93; P = 0.010), so it was lower than both categories below it; with 4.5–<5.0 mmol/L as the reference the ≥5.0 mmol/L contrast was 1.23 (0.90–1.69; P = 0.200), so it is not separated from the category above it (Supplementary Table S29).

**Table 2.** Composite and component outcomes by potassium response: adjusted hazard ratios.

|  | Events / N (rate per 1,000 py) | Adjusted HR (95% CI) | P |  | Events / N (rate per 1,000 py) | Adjusted HR (95% CI) | P |
|---|---|---|---|---|---|---|---|
| **Composite MI/stroke/death (n = 4,169; 623 events)** |  |  |  |  |  |  |  |
| **Achieved potassium (mmol/L)** |  |  |  | **ΔK□ (tertile)** |  |  |  |
| <4.0 | 213 / 1,360 (38.9) | 1.00 (reference) |  | T1 (lowest) | 247 / 1,485 (41.1) | 1.00 (reference) |  |
| 4.0–<4.5 | 236 / 1,612 (36.1) | 0.88 (0.73–1.07) | 0.191 | T2 | 184 / 1,370 (33.7) | 0.80 (0.66–0.98) | 0.032 |
| 4.5–<5.0 | 116 / 925 (31.1) | 0.65 (0.52–0.83) | <0.001 | T3 (highest) | 192 / 1,314 (35.1) | 0.75 (0.60–0.94) | 0.012 |
| ≥5.0 | 58 / 272 (47.7) | 0.80 (0.59–1.09) | 0.159 |  |  |  |  |
| **Incident heart failure (n = 3,632; 248 events)** |  |  |  |  |  |  |  |
| **Achieved potassium (mmol/L)** |  |  |  | **ΔK□ (tertile)** |  |  |  |
| <4.0 | 89 / 1,189 (17.7) | 1.00 (reference) |  | T1 (lowest) | 101 / 1,298 (18.2) | 1.00 (reference) |  |
| 4.0–<4.5 | 98 / 1,431 (16.3) | 0.83 (0.62–1.12) | 0.221 | T2 | 72 / 1,201 (14.5) | 0.73 (0.53–1.00) | 0.053 |
| 4.5–<5.0 | 38 / 790 (11.2) | 0.54 (0.36–0.80) | 0.002 | T3 (highest) | 75 / 1,133 (15.2) | 0.63 (0.44–0.89) | 0.009 |
| ≥5.0 | 23 / 222 (22.3) | 0.78 (0.48–1.27) | 0.321 |  |  |  |  |
| **All-cause death (n = 4,169; 379 events)** |  |  |  |  |  |  |  |
| **Achieved potassium (mmol/L)** |  |  |  | **ΔK□ (tertile)** |  |  |  |
| <4.0 | 129 / 1,360 (22.3) | 1.00 (reference) |  | T1 (lowest) | 154 / 1,485 (24.2) | 1.00 (reference) |  |
| 4.0–<4.5 | 131 / 1,612 (19) | 0.80 (0.62–1.03) | 0.085 | T2 | 114 / 1,370 (19.9) | 0.85 (0.66–1.10) | 0.224 |
| 4.5–<5.0 | 83 / 925 (21.2) | 0.75 (0.56–1.01) | 0.057 | T3 (highest) | 111 / 1,314 (19.2) | 0.74 (0.55–0.99) | 0.045 |
| ≥5.0 | 36 / 272 (28.2) | 0.80 (0.54–1.18) | 0.258 |  |  |  |  |
| **Myocardial infarction or stroke (n = 4,169; 319 events)** |  |  |  |  |  |  |  |
| **Achieved potassium (mmol/L)** |  |  |  | **ΔK□ (tertile)** |  |  |  |
| <4.0 | 112 / 1,360 (20.5) | 1.00 (reference) |  | T1 (lowest) | 132 / 1,485 (21.9) | 1.00 (reference) |  |
| 4.0–<4.5 | 133 / 1,612 (20.4) | 0.96 (0.74–1.25) | 0.772 | T2 | 91 / 1,370 (16.7) | 0.72 (0.54–0.96) | 0.024 |
| 4.5–<5.0 | 50 / 925 (13.4) | 0.56 (0.39–0.79) | 0.001 | T3 (highest) | 96 / 1,314 (17.6) | 0.68 (0.50–0.92) | 0.013 |
| ≥5.0 | 24 / 272 (19.7) | 0.68 (0.43–1.08) | 0.102 |  |  |  |  |
| **Incident heart failure (Fine-Gray) (n = 3,632; 248 events)** |  |  |  |  |  |  |  |
| **Achieved potassium (mmol/L)** |  |  |  | **ΔK□ (tertile)** |  |  |  |
| <4.0 | 89 / 1,189 (17.7) | 1.00 (reference) |  | T1 (lowest) | 101 / 1,298 (18.2) | 1.00 (reference) |  |
| 4.0–<4.5 | 98 / 1,431 (16.3) | 0.86 (0.64–1.15) | 0.299 | T2 | 72 / 1,201 (14.5) | 0.72 (0.53–0.99) | 0.041 |
| 4.5–<5.0 | 38 / 790 (11.2) | 0.58 (0.39–0.86) | 0.007 | T3 (highest) | 75 / 1,133 (15.2) | 0.68 (0.48–0.95) | 0.024 |
| ≥5.0 | 23 / 222 (22.3) | 0.89 (0.56–1.41) | 0.619 |  |  |  |  |
| **MI or stroke (Fine-Gray) (n = 4,169; 319 events)** |  |  |  |  |  |  |  |
| **Achieved potassium (mmol/L)** |  |  |  | **ΔK□ (tertile)** |  |  |  |
| <4.0 | 112 / 1,360 (20.5) | 1.00 (reference) |  | T1 (lowest) | 132 / 1,485 (21.9) | 1.00 (reference) |  |
| 4.0–<4.5 | 133 / 1,612 (20.4) | 1.03 (0.79–1.33) | 0.843 | T2 | 91 / 1,370 (16.7) | 0.74 (0.56–0.98) | 0.037 |
| 4.5–<5.0 | 50 / 925 (13.4) | 0.62 (0.43–0.87) | 0.007 | T3 (highest) | 96 / 1,314 (17.6) | 0.74 (0.55–1.00) | 0.047 |
| ≥5.0 | 24 / 272 (19.7) | 0.73 (0.47–1.13) | 0.159 |  |  |  |  |

Landmark day 180; the composite outcome is MI, stroke, or all-cause death; incident heart failure among those free of HF at baseline (Fine–Gray rows model the subdistribution hazard with death as a competing event). Reference categories: achieved K□ <4.0 mmol/L; ΔK□ tertile 1 (lowest). Adjusted for age, sex, race, MRA agent, baseline K□, baseline eGFR, hypertension, diabetes, CKD, atrial fibrillation, ischaemic heart disease, concomitant ACEi/ARB/thiazide/loop diuretic/potassium supplement; stratified by prior cerebrovascular disease and heart failure. Crude (unadjusted) HR for achieved K□ ≥5.0 vs <4.0, composite outcome, was 1.21 (0.91–1.62), attenuating to 0.80 (0.59–1.09) after adjustment; the sequence is given in Supplementary Table S30. ΔK□ tertiles are cut at ΔK□ = 0.10 and 0.60 mmol/L. Potassium is reported to one decimal place, so many patients sit exactly on a cut point and cannot be split between adjacent tertiles; the groups are therefore unequal (1,485 / 1,370 / 1,314). The incident heart-failure cohort is 4,169 − 511 with prevalent heart failure − 26 who acquired a first heart-failure code between the index date and the day-180 landmark = 3,632.

The change from baseline, which does not depend on these category boundaries, was associated with the composite outcome on a monotone gradient: 0.80 (95% CI, 0.66–0.98; P = 0.032) for the middle and 0.75 (0.60–0.94; P = 0.012) for the highest ΔK tertile, relative to the lowest (Table 2), reproduced with ΔK modelled continuously (Supplementary Figure S1). The two exposures derive from the same measurements and are reported together as complementary expressions of the early biochemical response rather than as independent analyses.

Crude estimates were closer to the null; the ≥5.0 mmol/L estimate moved from 1.21 to 0.80 after adjustment for age, race, eGFR and CKD (Supplementary Table S30).

### Time-Varying Association

The association was stronger in the first three years than thereafter (0.48; 95% CI, 0.34–0.69 versus 0.90; 0.64–1.26; P = 0.023 for the interaction) and also varied by calendar period of MRA initiation, which cannot be separated from follow-up duration in these data; the diagnostics are given in Supplementary Tables S4, S12–S14 and S25. The primary HRs are therefore reported as a summary association over follow-up rather than as an effect constant at every time point.

### Incident Heart Failure

Among the 3,632 patients free of HF at baseline and through the landmark, 248 incident HF and 227 competing deaths occurred. The same pattern appeared, more sharply. Relative to <4.0 mmol/L the adjusted HR was 0.54 (95% CI, 0.36–0.80; P = 0.002) for 4.5–<5.0 mmol/L, while neither 4.0–<4.5 mmol/L (0.83) nor ≥5.0 mmol/L (0.78) differed significantly from the reference (Table 2). As for the composite outcome, 4.5–<5.0 mmol/L was also lower than 4.0–<4.5 mmol/L (0.65; 0.44–0.95; P = 0.025) but not separated from ≥5.0 mmol/L (Supplementary Table S29). Fine–Gray subdistribution models treating death as a competing event gave the same shape (0.86, 0.58 and 0.89). For the change from baseline the gradient was monotone, with 0.63 (95% CI, 0.44–0.89; P = 0.009) for the highest ΔK tertile.

For this outcome the shape tests were mixed: of five comparisons only the spline given an explicit interior knot at 5.0 mmol/L improved on a linear term (P = 0.011), while the spline without it, and both fractional-polynomial comparisons, did not (Supplementary Table S26). None of these global tests locates a minimum. A departure from linearity near 5.0 mmol/L is therefore suggested for this outcome but not consistently supported.

### Absolute Risk

Covariate-standardised three-year cumulative incidence traced the same curve. For the composite outcome it was 11.31% (95% CI, 9.92–13.15), 10.13% (8.84–11.48), 7.78% (6.43–9.25) and 9.37% (7.51–11.79) across ascending categories; only 4.5–<5.0 mmol/L differed significantly from the reference, by 3.53 percentage points (1.71–5.77; Supplementary Table S19). For incident HF, risk fell from 4.44% to 2.61% at 4.5–<5.0 mmol/L, a difference of 1.84 percentage points (95% CI, 0.54–3.23).

### The Reference Category and Adjacent Contrasts

Because the reference category pools frank hypokalaemia with low-normal potassium, it was split at 3.5 mmol/L (Supplementary Table S29). Achieved potassium below 3.5 mmol/L did not differ significantly from 3.5–<4.0 mmol/L for either outcome. Restricted to 3.5 to <5.0 mmol/L (3,570 patients with 500 composite events; 3,126 with 197 heart-failure events), 4.5–<5.0 mmol/L gave 0.68 (0.53–0.88) and 0.54 (0.36–0.83) against 3.5–<4.0 mmol/L while 4.0–<4.5 mmol/L did not differ (0.91 and 0.86): the signal sits in the 4.5–<5.0 mmol/L band rather than in normalisation alone.

At the consensus threshold of 3.6 mmol/L, achieved potassium below the threshold was not associated with a significantly higher risk than potassium at or above it (composite outcome HR 1.22, 95% CI 0.96–1.56; incident HF 1.34, 0.91–1.96). With 3.6–<4.0 mmol/L as the reference, risk was again lowest at 4.5–<5.0 mmol/L (0.67, 0.52–0.86; HF 0.56, 0.36–0.85), whereas <3.6 mmol/L (1.06 and 1.11), 4.0–<4.5 mmol/L (0.90 and 0.86) and ≥5.0 mmol/L (0.82 and 0.81) did not differ significantly. Among 1,386 patients with a baseline potassium below 3.6 mmol/L, neither correction with supplementation (1.13, 0.80–1.60) nor failure to correct (1.03, 0.68–1.56) differed from correction without supplementation for the composite outcome; the heart-failure estimates were imprecise, with the highest point estimate for failure to correct (1.54, 0.84–2.84; Supplementary Table S35).

### Change in Potassium and Change in Renin

Plasma renin activity was paired to potassium in both protocol windows for 424 patients, of whom 346 also met the criteria for the landmark modelling cohort. The change in potassium correlated weakly with the change in renin and discriminated an unsuppressed renin at follow-up no better than chance.

### Subgroup and Sensitivity Analyses

Estimates for 4.5–<5.0 versus <4.0 mmol/L were directionally consistent across prespecified strata and analytic variations, ranging from 0.55 to 0.91, and no interaction test across patient subgroups reached significance (Figure 3; Supplementary Tables S16–S18). Among the 1,917 patients taking no potassium supplement, the group in which a potassium within the reference range corresponds to the PAMO definition of biochemical correction, the estimate was of similar magnitude to the overall estimate (0.68; 95% CI, 0.45–1.04; P for interaction with supplement use = 0.19), as it was among patients with no recorded work-up (0.73; 0.51–1.06); it was closest to the null among patients hypokalaemic at baseline (0.91; 0.55–1.50; P for interaction = 0.14). Estimates were also consistent across strata defined by the number of potassium measurements in the exposure window (Supplementary Table S20 and Supplementary Figure S2), and adjusting for treatment continuity left the ΔK estimates materially unchanged; because continuity is measured in the same window as the exposure, those analyses are reported in full in Supplementary Tables S6 and S24 rather than treated as confounding control.

**Figure 3.**
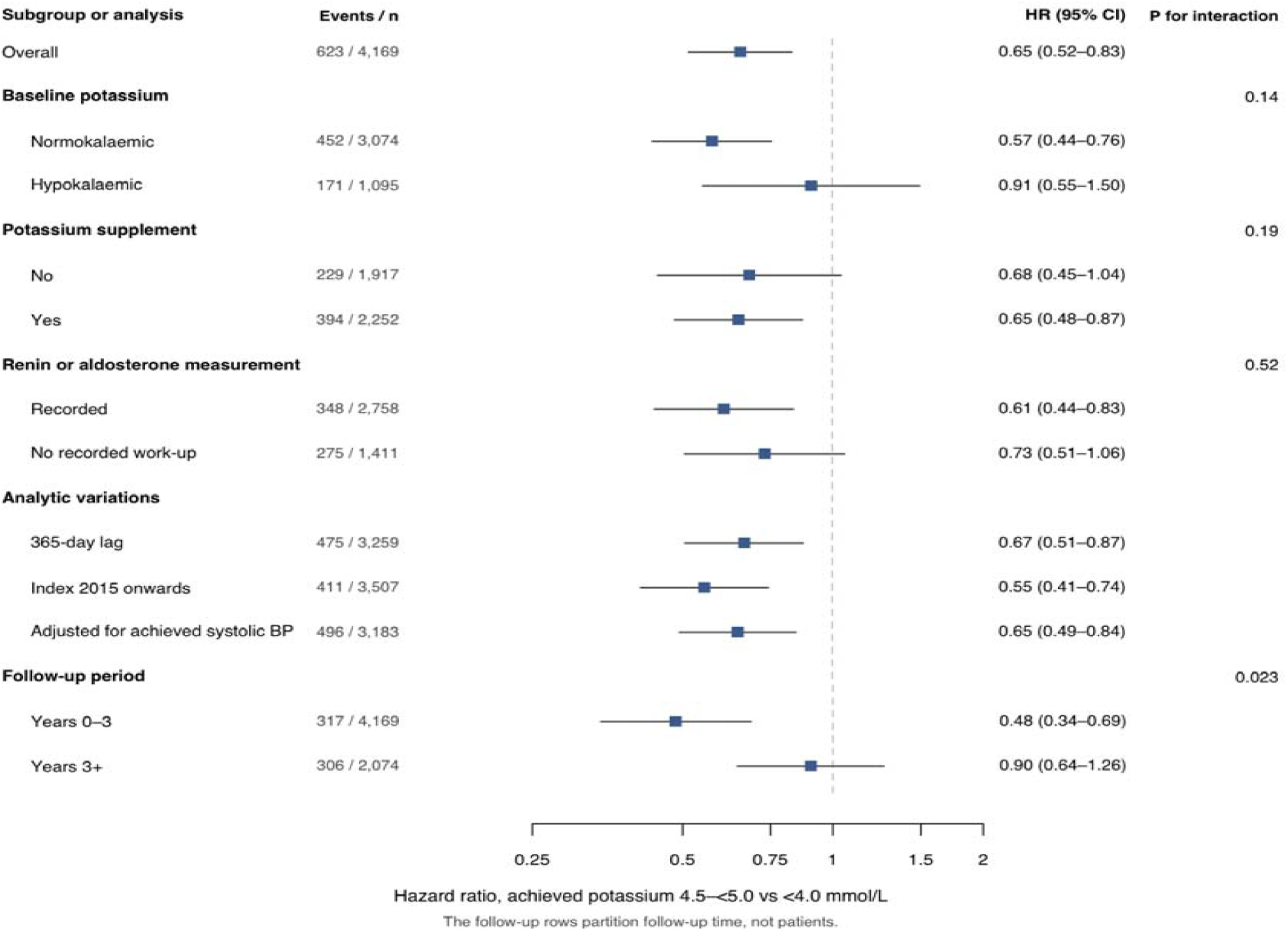
Sensitivity of the achieved-potassium association (4.5–<5.0 vs <4.0 mmol/L) with the composite outcome, across subgroups and analytic choices. “Years 0–3” and “Years 3+” under Follow-up period are not patient subgroups; they partition follow-up time within the same 4,169 patients. “No recorded work-up” = no renin or aldosterone measurement on record, which is not evidence that none was performed. Potassium supplement use is any potassium chloride, citrate or bicarbonate prescription from 180 days before MRA initiation to the day-180 landmark. P for interaction is from a likelihood-ratio test of the achieved-potassium category × subgroup term (3 df).

Patients excluded for missing paired potassium were healthier than those retained, but the 1,294 excluded after that step were substantially sicker (Supplementary Table S3). Inverse-probability-of-selection weighting did not materially change the estimate (Supplementary Table S22). Requiring two separately recorded PA diagnoses, or restricting to codes specific to PA, gave concordant composite estimates; the corresponding heart-failure estimates were attenuated and imprecise, reflecting fewer events in these subsets (Supplementary Tables S27, S28 and S33).

### Control Outcomes and Sensitivity to Unmeasured Confounding

The six negative control outcomes were treated as one family, and none was associated with the exposure after correction for multiple comparisons (Supplementary Tables S5 and S23). The positive control, achieved systolic BP, behaved as expected: higher achieved BP was associated with more composite events, and additional adjustment for it left the estimates essentially unchanged (Supplementary Table S11). E-values for the 4.5–<5.0 mmol/L contrast were 2.02 for the composite outcome and 2.44 for incident HF (Supplementary Table S15). Ascertaining outcomes at any encounter type rather than acute-care only left the composite estimates similar but attenuated the heart-failure estimates to include the null (Supplementary Table S32).

## Discussion

In 4,169 adults with PA starting an MRA, the potassium concentration attained in the first six months was associated with subsequent cardiovascular risk, and the association was not a simple gradient. Estimated risk was lowest among patients reaching 4.5–<5.0 mmol/L, significantly lower than in either band below it, while the estimate above 5.0 mmol/L was higher than at the minimum but not significantly so. A larger rise from baseline was associated with lower risk on a monotone gradient. The band associated with the lowest risk lies inside the reference range, and the association persisted when frankly hypokalaemic patients were removed.

These findings bear directly on how medical treatment of PA is monitored. Potassium is measured in nearly every treated patient, yet consensus criteria count correction of hypokalaemia, a potassium of at least 3.6 mmol/L, as an adequate response,^4^ and guidance sets no potassium target beyond normalisation.^7,11^ In this cohort that threshold did not separate patients by subsequent risk, whereas risk was lowest in a band almost 1 mmol/L higher. If confirmed, a potassium below 4.5 mmol/L under treatment would identify patients whose MRA dose, adherence or supplementation merits review, using a test that is already part of routine monitoring, and the potassium criterion in consensus definitions of treatment response would merit reconsideration. These data do not show that raising potassium lowers risk; they define the target that a titration trial should test.

### Potential Mechanisms

Several mechanisms, not mutually exclusive, could link the potassium attained under MRA therapy to subsequent risk. The first treats potassium as a read-out of receptor blockade in the kidney. Aldosterone acting through the mineralocorticoid receptor in the aldosterone-sensitive distal nephron increases sodium reabsorption through the epithelial sodium channel and, through the lumen-negative potential this generates, potassium secretion through ROMK and BK channels;^16^ an MRA reverses both, so the rise in potassium after initiation indexes the extent to which renal receptor activity has been blocked. The same receptor mediates the pro-fibrotic and pro-inflammatory actions of aldosterone in the heart and vasculature,^7^ and persistent renin suppression under treatment, which marks incomplete blockade, carries the excess cardiovascular risk of medically treated PA.^2,6^ The weak correlation between the change in potassium and the change in renin argues against treating the two as interchangeable, however: normokalaemia is reached at doses that leave renin suppressed in a substantial proportion of patients,^17^ plausibly because the tubular response to receptor blockade is also shaped by potassium supplementation, dietary intake, renal function and concomitant renin–angiotensin blockade, whereas renin reflects the restoration of sodium and volume balance.

The second is a direct effect of the potassium concentration itself. Low and low-normal potassium is arrhythmogenic,^18^ and in hypertensive, heart-failure and general populations the association between serum potassium and mortality is U-shaped, with its nadir between about 4.0 and 4.5 mmol/L.^12–15^ In experimental hypermineralocorticoid states cardiac fibrosis develops independently of potassium depletion,^19^ so a potassium-mediated pathway is more plausibly electrophysiological than structural; it would operate whether or not the concentration was reached with supplementation.

The third is that achieved potassium marks disease severity rather than mediating risk. Hypokalaemic PA is characterised by higher aldosterone, more suppressed renin and more target-organ damage than normokalaemic PA,^3^ so patients whose potassium remains below 4.5 mmol/L under treatment may have more severe or less responsive disease. Two observations bear on this. The association was, if anything, stronger among patients normokalaemic at baseline (HR 0.57) than among those hypokalaemic at baseline (0.91), and it persisted among patients taking no potassium supplement, the group in which correction of hypokalaemia corresponds to the PAMO definition of biochemical response,^4^ with no evidence of interaction by supplement use (Figure 3). Both suggest that the prognostic information is carried by the concentration attained rather than only by the severity of the pre-treatment deficit or by how the concentration was reached—relevant to routine care, where about half of treated patients receive supplements—but these data cannot separate marker from mediator. Finally, values of 5.0 mmol/L or more accompanied lower eGFR and more CKD and diabetes, and the crude estimate for that category moved from 1.21 to 0.80 with adjustment for renal function, so any upward limb is likely to reflect in part the renal impairment that both raises potassium and constrains MRA dosing,^20^ rather than an effect of potassium itself.

### A Minimum Inside the Reference Range

Serum potassium in PA is treated as a safety parameter: it confirms that hypokalaemia has been corrected and that hyperkalaemia has not supervened. If its prognostic information were exhausted by that role, the association would have disappeared once frankly hypokalaemic patients were excluded. It did not: within 3.5 to <5.0 mmol/L, the 4.5–<5.0 mmol/L band carried a HR of 0.68 (95% CI, 0.53–0.88) for the composite outcome and 0.54 (0.36–0.83) for incident HF, and it was also lower than the adjacent 4.0–<4.5 mmol/L band (0.74 and 0.65). What the data locate is a band inside the reference range, not a gradient, and the upper end is where the evidence stops.

This bears on how an adequate potassium response to medical treatment is defined. The threshold of 3.6 mmol/L shared by the PAMO and PASO definitions was agreed by Delphi panels to classify whether hypokalaemia had been corrected,^4,5^ rather than derived from outcome data; the PASO investigators noted that their criteria could not be validated against hard endpoints.^5^ In these data the threshold did not clearly separate patients by subsequent risk, and two features may explain this. First, the group above it is heterogeneous: risk at 4.0–<4.5 mmol/L did not differ significantly from that at 3.6–<4.0 mmol/L and was lowest only at 4.5–<5.0 mmol/L, so a single cut at 3.6 mmol/L averages bands with different risk. Second, the threshold is readily reached: potassium usually normalises within days of starting an MRA,^7^ and in one long-term cohort 90% of patients became normokalaemic without supplementation, whereas 48% reached a high-normal potassium of at least 4.3 mmol/L, a level some clinicians already target.^17^ Outside PA, the lowest mortality lies near 4.2 mmol/L.^11^ Reaching 3.6 mmol/L therefore marks the resolution of hypokalaemia rather than attainment of the band associated with the lowest risk. Whether the potassium criterion for an adequate response should be set higher cannot be established from an observational analysis, and the subgroup hypokalaemic at baseline was too small to exclude a difference, particularly for heart failure.

### Comparison With Potassium Ranges Derived From Other Populations

Outside PA the relationship between serum potassium and mortality is U-shaped, with both hypokalaemia and hyperkalaemia associated with excess mortality. The category with lowest risk here matches that reported in a cohort of 21,334 patients with incident HF, where 4.5–<5.0 mmol/L was likewise the reference band.^13^ The agreement is in which band carries the lowest risk, not in the shape of a curve, which these data do not resolve; the categories are wide and the formal shape tests were weak, so the correspondence should not be over-read.

### Change in Potassium Does Not Substitute for Renin

Renin, followed in only a minority of patients here as elsewhere, is examined next. Among 424 patients with plasma renin activity paired to potassium in the protocol windows, of whom 346 also met the criteria for the landmark modelling cohort, the change in potassium correlated weakly with the change in renin (Spearman rho, 0.15) and discriminated an unsuppressed renin at follow-up no better than chance (area under the curve, 0.523). In an independent cohort of 155 patients on long-term medical therapy, 90% became normokalaemic, yet among the 57 with renin measured, classification of biochemical control by potassium and by renin agreed in only 49.1%.^17^

A change in potassium therefore cannot be used as a proxy for renin status. Whether the two reflect different biological aspects of mineralocorticoid blockade is a further claim these data do not establish; weak correlation is equally consistent with measurement error. Nor can we claim that potassium carries information incremental to renin, since the two were never entered into a common model and the paired sample is too small to support that comparison.

### Incident Heart Failure

HF was examined as a co-primary outcome; the contrast at 4.5–<5.0 mmol/L was largest for this outcome, and Fine–Gray models allowing for competing death reproduced it. Receptor activation and the adequacy of blockade were not measured, so a lower heart-failure risk in one potassium band does not demonstrate that blockade was more complete in it.

### Strengths and Limitations

A landmark design measured the exposure before follow-up began,^21^ avoiding outcomes that preceded the exposure. Potassium was identified by LOINC code, not description matching. Death, competing with every non-fatal outcome, was handled by Fine–Gray models and standardisation of the subdistribution. A positive control performed as expected, and none of six negative controls was associated with the exposure after correction for multiple comparisons. E-values of 2.02 and 2.44 indicate the confounding needed to explain the primary associations;^22^ this does not establish that such confounding exists.

Some limitations qualify these findings. The case definition is administrative; the composite estimate was unchanged when restricted to codes specific to PA, but the heart-failure subsets were then too small to evaluate. MRA dose, titration, adherence, supplement dose and dietary intake were unknown, so the achieved potassium may reflect treatment exposure as well as response to it, and interaction tests by supplement use had limited power. The exposure is a single routine-care value; plasma and serum potassium differ systematically, which would preferentially affect the highest category. The categories are 0.5 mmol/L wide, so these data identify the category with the lowest estimated risk, not the concentration at which risk is lowest, and the upward limb above 5.0 mmol/L is suggested rather than established. The association varied by calendar period, which cannot be separated from follow-up duration; selection occurred in two steps and the weighted analysis is exploratory; the network is predominantly US-based. Finally, the exposure is attained rather than assigned, so these estimates do not establish that altering potassium alters risk.

## Conclusions

In adults with PA treated with an MRA, the potassium attained in the first six months was associated with subsequent cardiovascular risk and with incident HF with the lowest estimated risk in the 4.5–<5.0 mmol/L band, significantly lower than in either band below it. That band lies inside the reference range, and the association was not attributable to the correction of frank hypokalaemia alone. The band also lies well above the 3.6 mmol/L that consensus criteria accept as correction of hypokalaemia, so correction alone may be an insufficient treatment target. Whether the early potassium response can serve as a treatment target, rather than a marker of risk, requires prospective evaluation. Any such evaluation would need to compare titration strategies rather than attained concentrations, within the constraints that hyperkalaemia and renal function place on MRA dosing.

## Research in context

### Evidence before this study

We searched PubMed from database inception to July 2026, without language restriction, for studies using the terms (“primary aldosteronism” OR hyperaldosteronism) AND (“mineralocorticoid receptor antagonist” OR spironolactone OR eplerenone) AND potassium AND (cardiovascular OR “heart failure” OR mortality). Medically treated primary aldosteronism carries an excess of cardiovascular events over essential hypertension that is concentrated in patients whose renin remains suppressed, and a 2025 meta-analysis found unsuppressed post-treatment renin associated with fewer cardiovascular events. The PAMO consensus defines a complete biochemical response as correction of hypokalaemia (a potassium of at least 3.6 mmol/L, set by expert agreement without outcome data) without supplementation together with normalisation of renin, and the 2025 Endocrine Society guideline recommends monitoring potassium and renin to guide titration, but neither specifies a potassium target. Target ranges for serum potassium derive from hypertensive and heart-failure populations, in which the potassium–mortality association is U-shaped with a nadir near 4.0–4.5 mmol/L. We found no study relating the potassium concentration attained after MRA initiation to long-term cardiovascular outcomes in primary aldosteronism.

### Added value of this study

In 4,169 adults with primary aldosteronism in a multicentre electronic-health-record network, the potassium attained 14–180 days after MRA initiation was associated with myocardial infarction, stroke or death and with incident heart failure over a median of 3 years. Estimated risk was lowest at 4.5–<5.0 mmol/L—about a third lower than at <4.0 mmol/L and a quarter lower than at 4.0–<4.5 mmol/L—a band inside the reference range; the association persisted after excluding frankly hypokalaemic values and among patients taking no potassium supplement, and a larger rise from baseline was associated with lower risk. The change in potassium correlated only weakly with the change in renin, so potassium does not substitute for renin as a marker of treatment response. The consensus threshold for correction of hypokalaemia, 3.6 mmol/L, did not separate patients by subsequent risk.

### Implications of all the available evidence

In MRA-treated primary aldosteronism, an achieved potassium below 4.5 mmol/L, including low-normal values, identifies patients at higher subsequent cardiovascular and heart-failure risk from a test already performed in nearly every treated patient. Correction of hypokalaemia, the potassium criterion in current consensus definitions of treatment response, may therefore be an insufficient target, and that criterion merits reconsideration when those definitions are revised. Because potassium and renin dissociate, both should be monitored, as current guidance recommends. Whether titrating MRA therapy to reach 4.5–<5.0 mmol/L reduces risk, within the constraints that hyperkalaemia and renal function place on dosing, requires a prospective trial of a defined titration strategy.

## Supporting information

All Supplements

## Data Availability

The patient-level data underlying this study were accessed through the TriNetX Research Network and cannot be shared owing to the terms of the data use agreement governing access to the network.

## Contributors

SHC: conceptualisation, data curation, formal analysis, investigation, methodology, resources, software, validation, visualisation, writing – original draft, writing – review & editing

YHH: conceptualisation, investigation, methodology, writing – review & editing

YHL: data curation, investigation, methodology, project administration, writing – review & editing

SMY: investigation, methodology, project administration, resources, writing – review & editing

CJH: investigation, methodology, project administration, validation, writing – review & editing

YKP: data curation, project administration, resources, software, validation, writing – review & editing

YYC: data curation, resources, software, supervision, validation, writing – review & editing

HMC: conceptualisation, funding acquisition, investigation, methodology, resources, supervision, validation, visualisation, writing – original draft, writing – review & editing

All authors were involved in data interpretation, review, and writing of the manuscript.

## Data sharing

Statistical code is available from the corresponding author upon reasonable request. The patient-level data underlying this study were accessed through the TriNetX Research Network and cannot be shared owing to the terms of the data use agreement governing access to the network.

## Acknowledgments

None.

