## Supplementary material for "Early potassium response to mineralocorticoid receptor antagonist therapy and cardiovascular outcomes in primary aldosteronism: a landmark cohort study": All Supplements

**Contents**

**Supplementary Methods**

**Figure S1.** Change in potassium, modelled continuously (restricted cubic spline)

**Figure S2.** Timing of potassium measurements relative to the index date

**Figure S3.** Achieved potassium, modelled continuously (restricted cubic spline)

**Table S1.** RECORD reporting checklist

**Table S2.** Diagnosis, laboratory, and medication codes used to define the cohort, exposures, covariates, and outcomes

**Table S3.** Baseline characteristics of patients excluded for missing paired potassium or lost before the landmark, versus those retained in the analytic cohort

**Table S4.** Achieved potassium and the composite outcome, before and after 3 years of follow-up from the landmark

**Table S5.** Negative control outcomes regressed on the ΔK⁺ tertile exposure

**Table S6.** Sensitivity to treatment continuity: MRA fill count and proportion of days covered

**Table S7.** Attrition from any renin-related record to the paired plasma-renin-activity bridging set

**Table S8.** Spearman correlation between ΔK⁺ and Δplasma renin activity, by renin-window definition and restriction

**Table S9.** Additional renin discrimination statistics

**Table S10.** Systolic blood pressure measurement counts and reliability, last-value versus mean-based definitions

**Table S11.** Cox models for the blood-pressure positive control, and for potassium adjusted for blood pressure

**Table S12.** Achieved potassium and the composite outcome, by calendar period of MRA initiation and by follow-up era

**Table S13.** Descriptive statistics and interaction tests for calendar period

**Table S14.** Calendar period versus follow-up epoch: disentangling test (n = 4,169)

**Table S15.** E-value for unmeasured confounding of the primary achieved-potassium association

**Table S16.** Achieved K⁺ and ΔK⁺ by whether the patient was biochemically worked up for PA, with the formal interaction test

**Table S17.** Achieved K⁺ ≥5.0 mmol/L, by baseline eGFR, with the formal interaction test

**Table S18.** Achieved K⁺ and the composite outcome, by MRA agent, with the formal interaction test

**Table S19.** Covariate-standardised absolute risk and risk difference, 3 years from the landmark

**Table S20.** Timing of the exposure-defining potassium measurement, and the association by number of in-window measurements

**Table S21.** Tests for a non-linear or J-shaped association with achieved potassium

**Table S22.** Selection into the analytic cohort: standardised differences and inverse-probability-of-selection weighting

**Table S23.** Negative control outcomes ascertained on the primary outcome's pathway

**Table S24.** Change in potassium and the composite outcome, stratified by MRA fill count

**Table S25.** Proportional-hazards test for the incident-heart-failure model

**Table S26.** Tests for a non-linear association with achieved potassium and incident heart failure

**Table S27.** Composition of the cohort by hyperaldosteronism diagnosis code

**Table S28.** Primary models under a stricter diagnosis requirement

**Table S29.** The reference category split, the normal range alone, and contrasts between adjacent categories

**Table S30.** Sequential adjustment

**Table S31.** Race by achieved potassium, and the primary models estimated with and without race

**Table S32.** Outcome ascertainment: acute-care encounters only versus any encounter type

**Table S33.** Primary models restricted to patients carrying a diagnosis code specific to primary aldosteronism

**Table S34.** Sensitivity to adjustment for the concurrent change in eGFR

**Table S35.** The PAMO potassium threshold of 3.6 mmol/L

**Supplementary Methods**

Specimen types and measurement timing

Potassium was identified by Logical Observation Identifiers Names and Codes 2823-3 rather than by description matching. Plasma and serum potassium differ systematically, and haemolysed specimens are spuriously elevated; both would preferentially contaminate the highest achieved-potassium category. The specimen types included are listed in Supplementary Table S2. Because achieved potassium was defined, by construction, as the last value in the exposure window, the exposure-defining measurement fell systematically later than a typical in-window measurement (Supplementary Figure S2); the association was therefore examined stratified by the number of in-window measurements available.

Selection into the analytic cohort

Selection occurred in two steps of different character: availability of paired potassium measurements, then eligibility at the day-180 landmark. Both steps are characterised separately in Supplementary Table S3. Inverse-probability-of-selection weights were estimated within the second step and are reported as an exploratory reweighting (Supplementary Table S22), since several of the exclusions contributing to it are eligibility criteria rather than missing data.

Shape-of-association testing

Fractional polynomials were fitted by the Royston-Altman function-selection procedure: for each candidate power, and for each pair of powers, models were fitted over the conventional power set (-2, -1, -0.5, 0, 0.5, 1, 2, 3), and the best one-term and two-term fits were chosen by deviance and compared with a linear term on one and two degrees of freedom respectively (Supplementary Table S21).

Pre-specified robustness analyses

Robustness was examined in the following 12 pre-specified subgroups and analytic variations: baseline normokalaemia versus hypokalaemia; no potassium supplement; biochemical work-up recorded versus not recorded; strata of baseline eGFR; MRA agent (spironolactone versus eplerenone); index year from 2015 onward; spironolactone alone; adjustment for the concurrent change in eGFR; inverse-probability-of-selection weighting; a stricter two-diagnosis-code requirement for the case definition (evaluated because two separately dated codes raise diagnostic reproducibility without necessarily raising specificity); restriction to codes specific to PA; and treatment continuity (medication possession ratio). Results for each are given in Supplementary Tables S16-S18, S20, S22, S24 and S27-S28.

**Supplementary Figure S1. Change in potassium, modelled continuously (restricted cubic spline)**

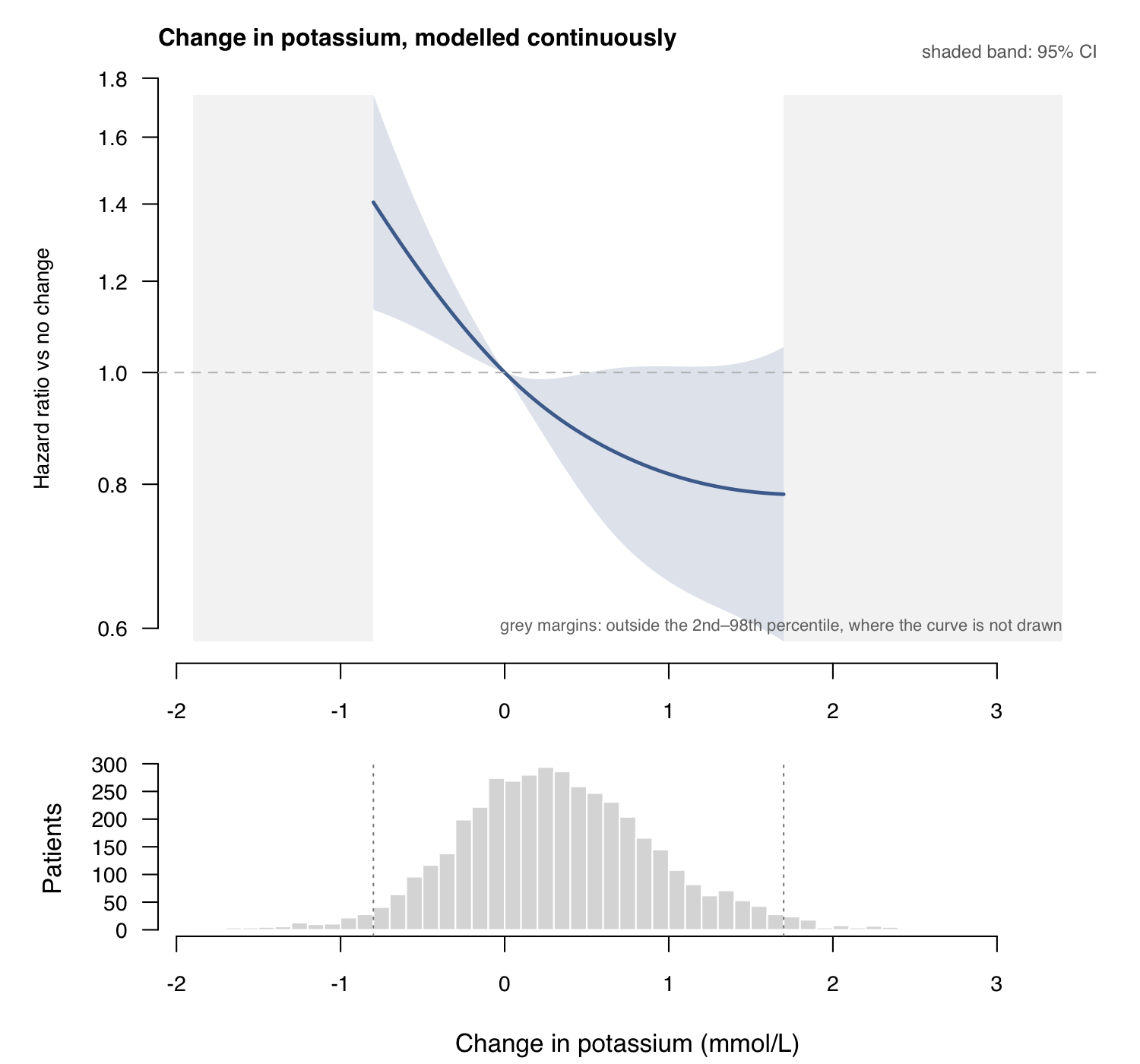

Reference = no change in potassium; shaded band is the 95% CI for the contrast of the spline basis at each value against the basis at the reference, computed by the delta method using the full covariance matrix of the spline coefficients (so the band correctly collapses to zero width at the reference). Natural cubic spline with 3 df; interior knots at ΔK⁺ = 0.1 and 0.6 mmol/L, boundary knots at the observed range. The curve is drawn over the 2nd–98th percentile only; the shaded margins of the panel mark where it stops, and both panels share a common x axis. Nested likelihood-ratio test: any association with ΔK⁺ P = 0.0043; non-linearity (spline vs. linear) P = 0.073. Histogram shows the patient distribution of ΔK⁺.

**Supplementary Figure S2. Timing of potassium measurements relative to the index date**

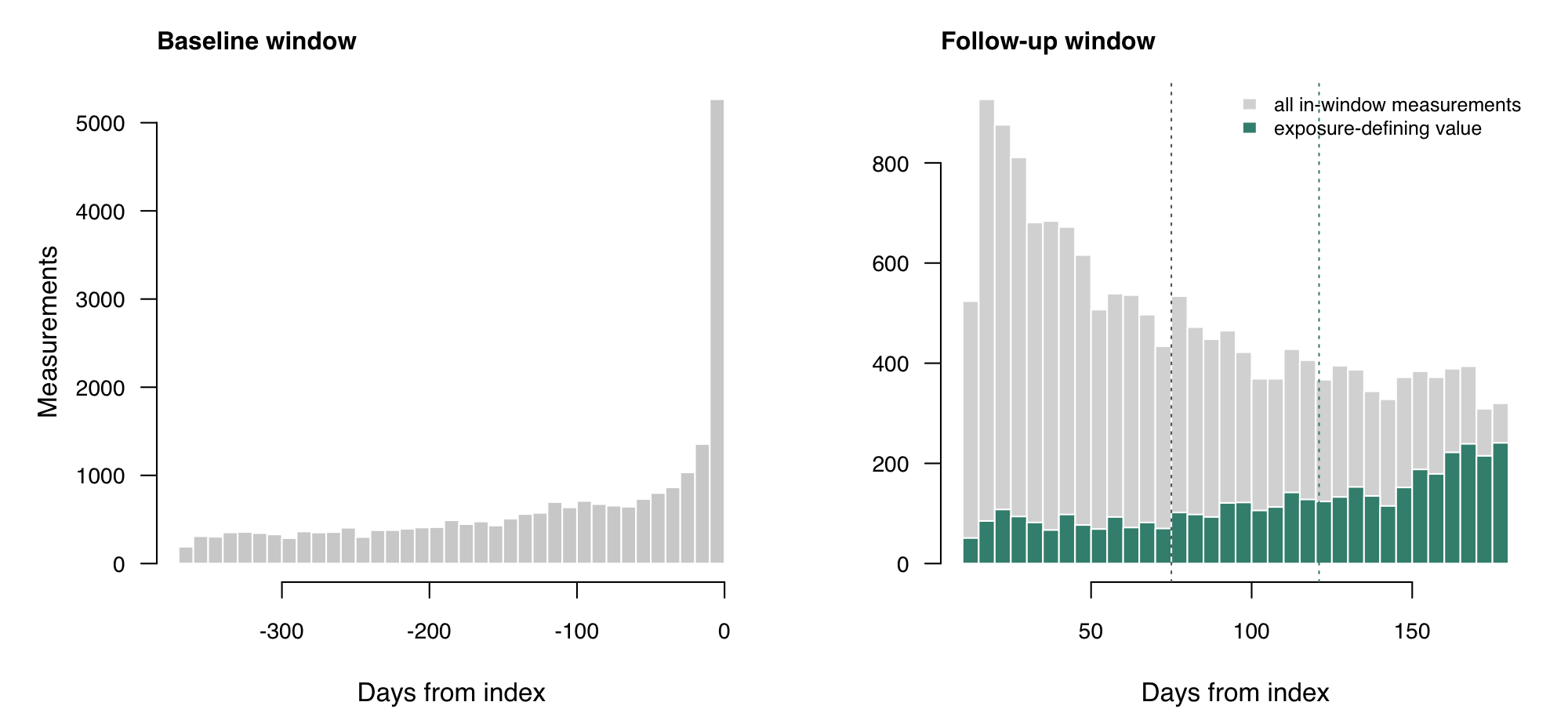

23,792 baseline and 16,578 follow-up potassium measurements in the analytic cohort; median day of the exposure-defining measurement (the Last value in days 14–180), day 121 (IQR 75–157); median 2 readings per patient in the follow-up window. The follow-up panel shows both distributions: every in-window measurement in grey (n = 16,578, median day 75) and, in colour, the one value per patient that defines the exposure (n = 4,169, median day 121). Dotted lines mark the two medians. Being the last value in the window, the exposure-defining measurement falls systematically later than a typical one. Stratifying by the number of in-window measurements does not change the association (P for interaction 0.64).

**Supplementary Figure S3. Achieved potassium, modelled continuously (restricted cubic spline)**

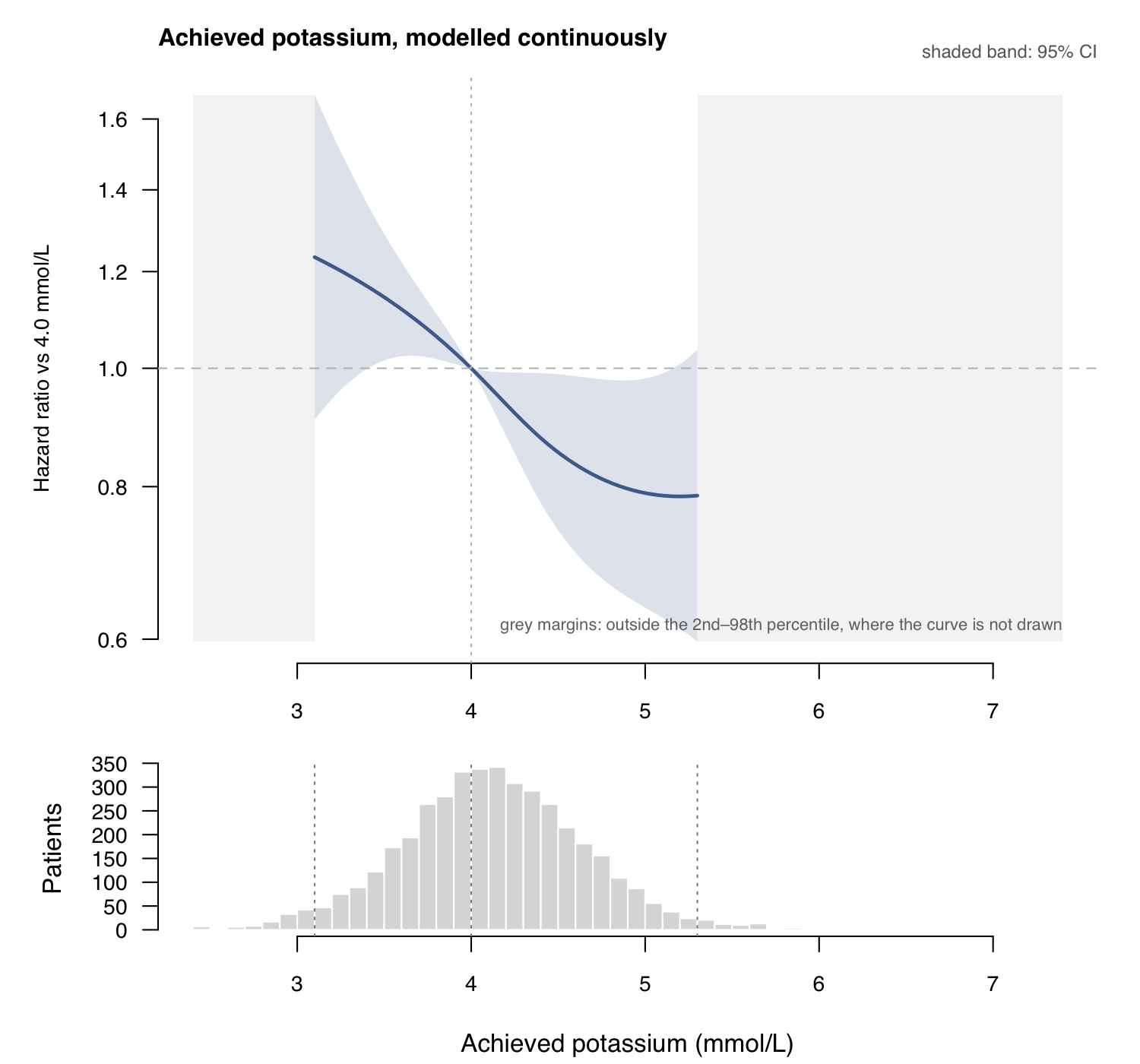

Reference = 4.0 mmol/L (the <4.0/4.0-4.5 category boundary used throughout). Shaded band is the 95% CI. Nested likelihood-ratio test: any association with achieved K⁺ P = 0.024; non-linearity (spline vs. linear) P = 0.62 (unchanged in direction; see the caveat below) (a restricted cubic spline is constrained to be linear beyond its outer knots, so a reversal confined to the sparse upper tail could not be expressed by this model) — no statistical evidence of a ceiling or reversal near 5.0 mmol/L; the attenuated, wide-CI point estimate for the achieved K⁺ ≥5.0 category in Table 2 is more consistent with sampling noise in a thin category (58 events) than with a real change in direction. Histogram shows the patient distribution of achieved K⁺. These two tests, and the fractional-polynomial and extra-knot alternatives, are collected in Supplementary Table S21.

**Supplementary Table S1.** RECORD reporting checklist

|  | **Item No.** | **STROBE items** | **Location in manuscript where items are reported** | **RECORD items** | **Location in manuscript where items are reported** |
| --- | --- | --- | --- | --- | --- |
| **Title and abstract** | | | | | |
|  | 1 | (a) Indicate the study’s design with a commonly used term in the title or the abstract (b) Provide in the abstract an informative and balanced summary of what was done and what was found | 1, 2 | RECORD 1.1: The type of data used should be specified in the title or abstract. When possible, the name of the databases used should be included.  RECORD 1.2: If applicable, the geographic region and timeframe within which the study took place should be reported in the title or abstract.  RECORD 1.3: If linkage between databases was conducted for the study, this should be clearly stated in the title or abstract. | 2  2  NA |
| **Introduction** | | | | | |
| Background rationale | 2 | Explain the scientific background and rationale for the investigation being reported | 3 |  |  |
| Objectives | 3 | State specific objectives, including any prespecified hypotheses | 4 |  |  |
| **Methods** | | | | | |
| Study Design | 4 | Present key elements of study design early in the paper | 5 |  |  |
| Setting | 5 | Describe the setting, locations, and relevant dates, including periods of recruitment, exposure, follow-up, and data collection | 5 |  |  |
| Participants | 6 | *(a) Cohort study* - Give the eligibility criteria, and the sources and methods of selection of participants. Describe methods of follow-up  *Case-control study* - Give the eligibility criteria, and the sources and methods of case ascertainment and control selection. Give the rationale for the choice of cases and controls  *Cross-sectional study* - Give the eligibility criteria, and the sources and methods of selection of participants  *(b) Cohort study* - For matched studies, give matching criteria and number of exposed and unexposed  *Case-control study* - For matched studies, give matching criteria and the number of controls per case | 5  NA | RECORD 6.1: The methods of study population selection (such as codes or algorithms used to identify subjects) should be listed in detail. If this is not possible, an explanation should be provided.  RECORD 6.2: Any validation studies of the codes or algorithms used to select the population should be referenced. If validation was conducted for this study and not published elsewhere, detailed methods and results should be provided.  RECORD 6.3: If the study involved linkage of databases, consider use of a flow diagram or other graphical display to demonstrate the data linkage process, including the number of individuals with linked data at each stage. | 5  NA  NA |
| Variables | 7 | Clearly define all outcomes, exposures, predictors, potential confounders, and effect modifiers. Give diagnostic criteria, if applicable. | 5–6 | RECORD 7.1: A complete list of codes and algorithms used to classify exposures, outcomes, confounders, and effect modifiers should be provided. If these cannot be reported, an explanation should be provided. | 5–6, Supplementary |
| Data sources/ measurement | 8 | For each variable of interest, give sources of data and details of methods of assessment (measurement).  Describe comparability of assessment methods if there is more than one group | 6, Supplementary |  |  |
| Bias | 9 | Describe any efforts to address potential sources of bias | 6–7 |  |  |
| Study size | 10 | Explain how the study size was arrived at | 8 |  |  |
| Quantitative variables | 11 | Explain how quantitative variables were handled in the analyses. If applicable, describe which groupings were chosen, and why | 6–7 |  |  |
| Statistical methods | 12 | (a) Describe all statistical methods, including those used to control for confounding  (b) Describe any methods used to examine subgroups and interactions  (c) Explain how missing data were addressed  (d) *Cohort study* - If applicable, explain how loss to follow-up was addressed  *Case-control study* - If applicable, explain how matching of cases and controls was addressed  *Cross-sectional study* - If applicable, describe analytical methods taking account of sampling strategy  (e) Describe any sensitivity analyses | 6–7 |  |  |
| Data access and cleaning methods |  | .. |  | RECORD 12.1: Authors should describe the extent to which the investigators had access to the database population used to create the study population.  RECORD 12.2: Authors should provide information on the data cleaning methods used in the study. | 6 |
| Linkage |  | .. |  | RECORD 12.3: State whether the study included person-level, institutional-level, or other data linkage across two or more databases. The methods of linkage and methods of linkage quality evaluation should be provided. | NA |
| **Results** | | | | | |
| Participants | 13 | (a) Report the numbers of individuals at each stage of the study (*e.g.*, numbers potentially eligible, examined for eligibility, confirmed eligible, included in the study, completing follow-up, and analysed)  (b) Give reasons for non-participation at each stage.  (c) Consider use of a flow diagram | 8 | RECORD 13.1: Describe in detail the selection of the persons included in the study (*i.e.,* study population selection) including filtering based on data quality, data availability and linkage. The selection of included persons can be described in the text and/or by means of the study flow diagram. | 8 |
| Descriptive data | 14 | (a) Give characteristics of study participants (*e.g.*, demographic, clinical, social) and information on exposures and potential confounders  (b) Indicate the number of participants with missing data for each variable of interest  (c) *Cohort study* - summarise follow-up time (*e.g.*, average and total amount) | 8, Table 1 |  |  |
| Outcome data | 15 | *Cohort study* - Report numbers of outcome events or summary measures over time  *Case-control study* - Report numbers in each exposure category, or summary measures of exposure  *Cross-sectional study* - Report numbers of outcome events or summary measures | 8–11 |  |  |
| Main results | 16 | (a) Give unadjusted estimates and, if applicable, confounder-adjusted estimates and their precision (e.g., 95% confidence interval). Make clear which confounders were adjusted for and why they were included  (b) Report category boundaries when continuous variables were categorized  (c) If relevant, consider translating estimates of relative risk into absolute risk for a meaningful time period | 8–11 |  |  |
| Other analyses | 17 | Report other analyses done—e.g., analyses of subgroups and interactions, and sensitivity analyses | 10–11 |  |  |
| **Discussion** | | | | | |
| Key results | 18 | Summarise key results with reference to study objectives | 12 |  |  |
| Limitations | 19 | Discuss limitations of the study, taking into account sources of potential bias or imprecision. Discuss both direction and magnitude of any potential bias | 15 | RECORD 19.1: Discuss the implications of using data that were not created or collected to answer the specific research question(s). Include discussion of misclassification bias, unmeasured confounding, missing data, and changing eligibility over time, as they pertain to the study being reported. | 15 |
| Interpretation | 20 | Give a cautious overall interpretation of results considering objectives, limitations, multiplicity of analyses, results from similar studies, and other relevant evidence | 16 |  |  |
| Generalisability | 21 | Discuss the generalisability (external validity) of the study results | 15 |  |  |
| **Other Information** | | | | | |
| Funding | 22 | Give the source of funding and the role of the funders for the present study and, if applicable, for the original study on which the present article is based | 2 |  |  |
| Accessibility of protocol, raw data, and programming code |  |  |  | RECORD 22.1: Authors should provide information on how to access any supplemental information such as the study protocol, raw data, or programming code. |  |

**Supplementary Table S2. Diagnosis, laboratory, and medication codes used to define the cohort, exposures, covariates, and outcomes**

| **Domain** | **Concept** | **System** | **Definition** |
| --- | --- | --- | --- |
| Cohort | MRA exposure | RxNorm ingredient | spironolactone\|eplerenone |
| Exposure | Serum potassium | LOINC code | 2823-3 (serum or plasma), mmol/L only. Whole blood 6298-4, arterial 32713-0, venous 39789-3 and capillary 39790-1 are deliberately NOT admitted; a sensitivity analysis admitting them is reported and changes nothing. |
| Covariate | Serum creatinine | LOINC description | ^Creatinine \[Mass/volume\] in Serum or Plasma |
| Covariate | Systolic blood pressure | LOINC code | 8480-6, 76534-7, 8459-0, 8460-8, 8461-6 |
| Design | Acute-care encounter types | Encounter type | IMP, ACUTE, EMER |
| Comorbidity | htn | ICD-9/10-CM | ^I1[0-6] |
| Comorbidity | dm | ICD-9/10-CM | ^E1[01] |
| Comorbidity | ckd | ICD-9/10-CM | ^N18 |
| Comorbidity | hf | ICD-9/10-CM | ^I50 |
| Comorbidity | af | ICD-9/10-CM | ^I48 |
| Comorbidity | ihd | ICD-9/10-CM | ^I2[0-5] |
| Comorbidity | cvd | ICD-9/10-CM | ^I6[0-9] |
| Comorbidity | obes | ICD-9/10-CM | ^E66 |
| Comorbidity | esrd | ICD-9/10-CM | ^N18\.?6\|^Z99\.?2 |
| Outcome | mi | ICD-9/10-CM | ^I2[12] |
| Outcome | stroke | ICD-9/10-CM | ^I6[0-4] |
| Outcome | hf_ev | ICD-9/10-CM | ^I50 |
| Outcome | af_ev | ICD-9/10-CM | ^I48 |
| Negative control | cataract | ICD-9/10-CM | ^H25\|^H26 |
| Negative control | zoster | ICD-9/10-CM | ^B02 |
| Negative control | hernia | ICD-9/10-CM | ^K40 |
| Negative control | hipfx | ICD-9/10-CM | ^S72 |
| Medication | acei | RxNorm ingredient | lisinopril\|enalapril\|ramipril\|benazepril\|captopril\|quinapril\|fosinopril\|perindopril\|trandolapril\|moexipril |
| Medication | arb | RxNorm ingredient | losartan\|valsartan\|irbesartan\|candesartan\|olmesartan\|telmisartan\|azilsartan\|eprosartan |
| Medication | thiazide | RxNorm ingredient | hydrochlorothiazide\|chlorthalidone\|indapamide\|metolazone\|chlorothiazide |
| Medication | loop | RxNorm ingredient | furosemide\|bumetanide\|torsemide\|ethacrynic |
| Medication | ksupp | RxNorm ingredient | potassium chloride\|potassium citrate\|potassium bicarbonate |
| Medication | ccb | RxNorm ingredient | amlodipine\|nifedipine\|diltiazem\|verapamil\|felodipine\|nicardipine |
| Medication | bb | RxNorm ingredient | metoprolol\|atenolol\|carvedilol\|bisoprolol\|labetalol\|propranolol\|nebivolol |
| Cohort | Primary aldosteronism | ICD-10-CM | ^E26 (E26.9 conclusive-unspecified, E26.09, E26.81, E26.01, E26.89, E26.02, E26.1, E26.0). Applied by TriNetX at extraction as a loose ICD-based definition; no confirmatory-testing requirement. |
| Outcome | All-cause death | Vital status | death_date in the TriNetX patient table. Mortality capture is incomplete and varies by contributing site; MI or stroke is therefore presented as a secondary outcome (see Table S12-S14). |
| Note | Systolic blood pressure | — | Matched by LOINC code, not description text: the dominant code 8480-6 is described 'Blood Pressure Systolic', with the words reversed relative to the usual phrasing. Invasive arterial code 76215-3 was excluded. |

**Supplementary Table S3. Baseline characteristics of patients excluded for missing paired potassium or lost before the landmark, versus those retained in the analytic cohort**

| **Group** | **N** | **% with SBP** | **Age** | **Baseline K⁺** | **Baseline eGFR** | **Baseline SBP** | **HTN %** | **DM %** | **CKD %** | **HF %** | **AF %** | **IHD %** |
| --- | --- | --- | --- | --- | --- | --- | --- | --- | --- | --- | --- | --- |
| No paired potassium (excluded before landmark selection) | 4614 | 70.2 | 57 | 3.9 | 77.8 | 143.6 | 79 | 24 | 19 | 10 | 7 | 16 |
| In the analytic cohort | 4169 | 83.3 | 59 | 3.8 | 76 | 144.9 | 87 | 31 | 25 | 12 | 9 | 20 |
| Excluded after the paired-potassium step (ESRD/dialysis, event or loss before the day-180 landmark, or incomplete covariates) | 1294 | 86.6 | 58.3 | 3.8 | 67.2 | 144.5 | 91 | 38 | 41 | 26 | 15 | 34 |

**Supplementary Table S4. Achieved potassium and the composite outcome, before and after 3 years of follow-up from the landmark**

| **Model** | **Term** | **HR (95% CI)** | **P** |
| --- | --- | --- | --- |
| Composite: achieved K⁺ — follow-up years 0–3 | Achieved K⁺ 4.0–4.5 vs <4.0 mmol/L | 0.84 (0.65–1.09) | 0.191 |
|  | Achieved K⁺ 4.5–5.0 vs <4.0 mmol/L | 0.48 (0.34–0.69) | <0.001 |
|  | Achieved K⁺ ≥5.0 vs <4.0 mmol/L | 0.62 (0.40–0.98) | 0.039 |
| Composite: achieved K⁺ — follow-up years 3+ | Achieved K⁺ 4.0–4.5 vs <4.0 mmol/L | 0.96 (0.72–1.28) | 0.770 |
|  | Achieved K⁺ 4.5–5.0 vs <4.0 mmol/L | 0.90 (0.64–1.26) | 0.552 |
|  | Achieved K⁺ ≥5.0 vs <4.0 mmol/L | 1.05 (0.69–1.60) | 0.808 |

Time-varying association: the HR for achieved K⁺ 4.5–5.0 vs <4.0 is stronger in the first 3 years (0.48, 95% CI 0.34–0.69) than beyond 3 years (0.90, 95% CI 0.64–1.26).

**Supplementary Table S5. Negative control outcomes regressed on the ΔK⁺ tertile exposure**

| **Negative control** | **Term** | **HR (95% CI)** | **P** |
| --- | --- | --- | --- |
| negative control: cataract | ΔK⁺ tertile 2 vs tertile 1 (lowest) | 1.23 (0.99–1.52) | 0.067 |
|  | ΔK⁺ tertile 3 (highest) vs tertile 1 | 1.03 (0.80–1.32) | 0.815 |
| negative control: zoster | ΔK⁺ tertile 2 vs tertile 1 (lowest) | 0.73 (0.46–1.18) | 0.197 |
|  | ΔK⁺ tertile 3 (highest) vs tertile 1 | 0.75 (0.43–1.29) | 0.299 |
| negative control: hernia | ΔK⁺ tertile 2 vs tertile 1 (lowest) | 1.16 (0.69–1.94) | 0.572 |
|  | ΔK⁺ tertile 3 (highest) vs tertile 1 | 1.02 (0.57–1.81) | 0.954 |

Cataract, herpes zoster, and inguinal hernia: outcomes MR blockade cannot plausibly prevent. Treated as one family of six hypotheses, one test per control (highest versus lowest ΔK⁺ tertile), none is associated with the exposure after Benjamini–Hochberg correction; cellulitis has the smallest raw P value, 0.046, and an adjusted P of 0.274. Hip fracture (a frailty probe rather than a pure negative control) is reported in the text (HR 1.26, 95% CI 0.48–3.32) but omitted here for small numbers (45 events).

**Supplementary Table S6. Sensitivity to treatment continuity: MRA fill count and proportion of days covered**

| **Model** | **Term** | **HR (95% CI)** | **P** |
| --- | --- | --- | --- |
| Composite: ΔK⁺ + fill count | ΔK⁺ tertile 2 vs tertile 1 (lowest) | 0.80 (0.65–0.97) | 0.027 |
|  | ΔK⁺ tertile 3 (highest) vs tertile 1 | 0.74 (0.59–0.92) | 0.007 |
|  | MRA fills, 2–3 vs 1 | 1.21 (0.98–1.49) | 0.072 |
|  | MRA fills, 4–6 vs 1 | 1.31 (1.04–1.65) | 0.024 |
|  | MRA fills, ≥7 vs 1 | 1.40 (1.11–1.76) | 0.004 |
| Composite: ΔK⁺ — PDC subset, without pdc | ΔK⁺ tertile 2 vs tertile 1 (lowest) | 0.84 (0.58–1.24) | 0.383 |
|  | ΔK⁺ tertile 3 (highest) vs tertile 1 | 0.64 (0.41–0.99) | 0.044 |
| Composite: ΔK⁺ — PDC subset, with pdc | ΔK⁺ tertile 2 vs tertile 1 (lowest) | 0.85 (0.58–1.24) | 0.389 |
|  | ΔK⁺ tertile 3 (highest) vs tertile 1 | 0.64 (0.41–0.99) | 0.045 |
|  | Proportion of days covered | 0.96 (0.62–1.50) | 0.867 |

More fills was associated with higher, not lower, risk — the opposite direction to healthy-adherer bias. Fill count barely relates to ΔK⁺ (Spearman rho 0.06, P = 0.0003). Because fill count is measured in the same 14–180 day window as the exposure, adjusting for it is post-baseline adjustment; the finding is therefore reported as no evidence of healthy-adherer bias in the expected direction, not as positive evidence against it.

**Supplementary Table S7.** Attrition from any renin-related record to the paired plasma-renin-activity bridging set

| **Step** | **Patients remaining** | **Lost at this step** |
| --- | --- | --- |
| any record matching 'renin' (includes the ratio) | 4364 |  |
| true renin only (aldosterone/renin ratio removed) | 4346 | 18 |
| has >=2 renin values at any time | 2341 | 2005 |
| has >=1 pre AND >=1 post, any window | 1111 | 1230 |
| pre within 2y, post within 2y | 884 | 227 |
| pre -365..0, post 14-180 (protocol windows) | 462 | 422 |
| same assay unit at both timepoints | 459 | 3 |
| ...and the unit is ng/mL/h (plasma renin activity) | 451 | 8 |
| ...and paired potassium in the same windows | 424 | 27 |
| ...and a single MRA agent (bridging set used in S8/S9) | 424 | 0 |

**Supplementary Table S8. Spearman correlation between ΔK⁺ and Δplasma renin activity, by renin-window definition and restriction**

| Window definition | Restriction | N | Spearman rho |
| --- | --- | --- | --- |
| Protocol windows (pre -365 to 0 d, post 14-180 d) | all | 424 | 0.150 (0.050 to 0.247), P = 0.002 |
|  | no K supplement | 74 | 0.239 (-0.003 to 0.454), P = 0.040 |
|  | normokalaemic at baseline | 317 | 0.134 (0.018 to 0.247), P = 0.017 |
|  | both restrictions | 63 | 0.213 (-0.052 to 0.450), P = 0.094 |
| ±1 year | all | 584 | 0.114 (0.028 to 0.198), P = 0.006 |
|  | no K supplement | 101 | 0.142 (-0.067 to 0.339), P = 0.157 |
|  | normokalaemic at baseline | 440 | 0.156 (0.058 to 0.252), P = <0.001 |
|  | both restrictions | 88 | 0.145 (-0.079 to 0.355), P = 0.178 |
| ±2 years | all | 754 | 0.104 (0.028 to 0.178), P = 0.004 |
|  | no K supplement | 127 | 0.183 (-0.002 to 0.355), P = 0.040 |
|  | normokalaemic at baseline | 551 | 0.138 (0.051 to 0.224), P = 0.001 |
|  | both restrictions | 111 | 0.218 (0.022 to 0.399), P = 0.021 |
| Any timing | all | 1049 | 0.089 (0.025 to 0.152), P = 0.004 |
|  | no K supplement | 171 | 0.167 (0.009 to 0.318), P = 0.029 |
|  | normokalaemic at baseline | 767 | 0.136 (0.061 to 0.208), P = <0.001 |
|  | both restrictions | 155 | 0.190 (0.024 to 0.346), P = 0.018 |

**Supplementary Table S9. Additional renin discrimination statistics**

| Item | Value | Detail |
| --- | --- | --- |
| Wilcoxon P, dK by an unsuppressed renin at follow-up | 0.423 |  |
| discrimination of dK for an unsuppressed renin at follow-up (AUC) | 0.523 | 0.5 means no discrimination at all |

n = 424 with clean paired plasma renin activity at the protocol windows (pre −365..0 d, post 14–180 d, same assay unit, ng/mL/h). ΔK⁺ vs Δrenin: Spearman rho = 0.15 (P = 0.002) — real but weak, and AUC 0.523 for discriminating an unsuppressed renin at follow-up is indistinguishable from no discrimination. Report as null: ΔK⁺ should not be framed as a proxy for renin status. A suppressed pre-treatment value was not required for entry, so this describes renin status at follow-up rather than a transition. The threshold is a plasma renin activity of 1.0 ng/mL/h or greater; the PAMO consensus words the same threshold as greater than 1 ng/mL/h, which would move 8 of the 424 patients.

**Supplementary Table S10.** Systolic blood pressure measurement counts and reliability, last-value versus mean-based definitions

| **Item** | **Value** | **Detail** |
| --- | --- | --- |
| patients with baseline SBP | 8339 | denominator: source population with the relevant SBP measurements |
| patients with post-window SBP | 6646 | denominator: source population with the relevant SBP measurements |
| patients with paired SBP | 6288 | denominator: source population with the relevant SBP measurements |
| median readings per patient, baseline | 7 | denominator: source population with the relevant SBP measurements |
| median readings per patient, post window | 4 | denominator: source population with the relevant SBP measurements |
| IQR readings, post window | 2 – 10 | denominator: source population with the relevant SBP measurements |
| patients with >=3 readings in BOTH windows | 3667 | denominator: source population with the relevant SBP measurements |
| median within-person SD of in-window SBP | 11.3 | mmHg; denominator: source population with the relevant SBP measurements |
| Spearman rho, last-based vs mean-based dSBP | 0.605 | denominator: source population with the relevant SBP measurements |
| SD of dSBP, last-based | 24.6 | denominator: source population with the relevant SBP measurements |
| SD of dSBP, mean-based | 17.4 | a smaller SD is the noise coming out; denominator: source population with the relevant SBP measurements |
| reliability of mean-based dSBP | 0.458 | denominator: source population with the relevant SBP measurements |
| reliability of last-based dSBP | 0.002 | denominator: source population with the relevant SBP measurements |
| Spearman rho, dK vs mean-based dSBP | -0.191 | P = 2e-27; denominator: analytic cohort |
| achieved-SBP cohort | 3183 | denominator: analytic cohort |
| Spearman rho, achieved K vs achieved SBP | -0.112 | P = 2.57e-10; denominator: analytic cohort |

**Supplementary Table S11. Cox models for the blood-pressure positive control, and for potassium adjusted for blood pressure**

| **Model** | **Term** | **HR (95% CI)** | **P** |
| --- | --- | --- | --- |
| Composite: ΔSBP (last value) tertile | ΔSBP (last value) T2, middle vs T1 (greatest fall) | 0.95 (0.76–1.19) | 0.666 |
|  | ΔSBP (last value) T3, least fall vs T1 (greatest fall) | 0.99 (0.80–1.23) | 0.937 |
| Composite: ΔSBP (last value) per 10 mmHg fall | Per 10 mmHg fall | 1.00 (0.97–1.04) | 0.876 |
| Composite: ΔSBP (mean-based) tertile | ΔSBP (mean-based) T2, middle vs T1 (greatest fall) | 1.08 (0.87–1.34) | 0.486 |
|  | ΔSBP (mean-based) T3, least fall vs T1 (greatest fall) | 0.95 (0.76–1.19) | 0.655 |
| Composite: ΔSBP (mean-based) per 10 mmHg fall | Per 10 mmHg fall | 1.00 (0.95–1.06) | 0.860 |
| Composite: ΔSBP (median-based) tertile | ΔSBP (median-based) T2, middle vs T1 (greatest fall) | 1.04 (0.83–1.29) | 0.741 |
|  | ΔSBP (median-based) T3, least fall vs T1 (greatest fall) | 0.95 (0.76–1.18) | 0.630 |
| Composite: ΔSBP (median-based) per 10 mmHg fall | Per 10 mmHg fall | 1.00 (0.95–1.05) | 0.914 |
| Composite: ΔSBP (mean-based) tertile — ≥3 readings per window | ΔSBP (mean-based) T2, middle vs T1 (greatest fall) | 1.15 (0.89–1.48) | 0.286 |
|  | ΔSBP (mean-based) T3, least fall vs T1 (greatest fall) | 1.00 (0.77–1.30) | 0.985 |
| Composite: ΔSBP (mean-based) per 10 mmHg fall — ≥3 readings per window | Per 10 mmHg fall | 0.98 (0.92–1.05) | 0.517 |
| Composite: ΔK⁺ — SBP subset, no ΔSBP | ΔK⁺ tertile 2 vs tertile 1 (lowest) | 0.68 (0.54–0.85) | <0.001 |
|  | ΔK⁺ tertile 3 (highest) vs tertile 1 | 0.65 (0.51–0.84) | <0.001 |
| Composite: ΔK⁺ — adjusted for ΔSBP (mean-based) | ΔK⁺ tertile 2 vs tertile 1 (lowest) | 0.68 (0.54–0.85) | <0.001 |
|  | ΔK⁺ tertile 3 (highest) vs tertile 1 | 0.64 (0.50–0.83) | <0.001 |
|  | Mean-based ΔSBP, per mmHg | 1.00 (0.99–1.00) | 0.504 |
| Composite: achieved K⁺ — SBP subset, no ΔSBP | Achieved K⁺ 4.0–4.5 vs <4.0 mmol/L | 0.82 (0.67–1.02) | 0.073 |
|  | Achieved K⁺ 4.5–5.0 vs <4.0 mmol/L | 0.62 (0.47–0.81) | <0.001 |
|  | Achieved K⁺ ≥5.0 vs <4.0 mmol/L | 0.73 (0.52–1.04) | 0.084 |
| Composite: achieved K⁺ — adjusted for ΔSBP (mean-based) | Achieved K⁺ 4.0–4.5 vs <4.0 mmol/L | 0.82 (0.66–1.01) | 0.067 |
|  | Achieved K⁺ 4.5–5.0 vs <4.0 mmol/L | 0.61 (0.47–0.80) | <0.001 |
|  | Achieved K⁺ ≥5.0 vs <4.0 mmol/L | 0.72 (0.51–1.03) | 0.073 |
|  | Mean-based ΔSBP, per mmHg | 1.00 (0.99–1.00) | 0.526 |
| Composite: achieved SBP (positive control) | Achieved SBP 130–140 vs <130 mmHg | 1.11 (0.86–1.42) | 0.425 |
|  | Achieved SBP 140–160 vs <130 mmHg | 1.40 (1.12–1.76) | 0.004 |
|  | Achieved SBP ≥160 vs <130 mmHg | 1.44 (1.05–1.97) | 0.022 |
| Composite: achieved SBP per 10 mmHg | Achieved SBP, per 10 mmHg | 1.09 (1.04–1.15) | <0.001 |
| Composite: achieved K⁺ — adjusted for achieved SBP | Achieved K⁺ 4.0–4.5 vs <4.0 mmol/L | 0.84 (0.68–1.05) | 0.121 |
|  | Achieved K⁺ 4.5–5.0 vs <4.0 mmol/L | 0.65 (0.49–0.84) | 0.001 |
|  | Achieved K⁺ ≥5.0 vs <4.0 mmol/L | 0.77 (0.54–1.10) | 0.151 |

The ΔSBP analysis stays null even after averaging all in-window readings to reduce measurement noise (reliability of mean-based ΔSBP 0.458 vs 0.002 for the last-value definition; the subgroup with at least three readings in each window comprises 2,099 patients with 375 events — see Table S10); the achieved-SBP framing does show the expected gradient (140–160 vs <130 mmHg: HR 1.40; ≥160: HR 1.44). The potassium association is unchanged by adjustment for either ΔSBP or achieved SBP, and ΔK⁺ correlates only weakly with ΔSBP (Spearman rho −0.19). The positive control is the ACHIEVED systolic pressure, and it behaves as it should (1.09 per 10 mmHg, 1.04–1.15). The CHANGE in systolic pressure is not treated as a control: its reliability here is 0.458 for a mean-based and 0.002 for a last-value-based change, so its null result is uninformative about the analysis. The candidate explanations for that null are: the candidate explanations are measurement error, confounding by the indication for measuring blood pressure, and a short exposure window, and the same measurement concerns apply to ΔK⁺.

**Supplementary Table S12. Achieved potassium and the composite outcome, by calendar period of MRA initiation and by follow-up era**

| **Model** | **Term** | **HR (95% CI)** | **P** | **N** | **Events** |
| --- | --- | --- | --- | --- | --- |
| Composite: achieved K⁺ — index year ≤2009 | Achieved K⁺ 4.0–4.5 vs <4.0 mmol/L | 0.38 (0.15–0.99) | 0.049 | 101 | 42 |
|  | Achieved K⁺ 4.5–5.0 vs <4.0 mmol/L | 0.36 (0.12–1.11) | 0.075 | 101 | 42 |
|  | Achieved K⁺ ≥5.0 vs <4.0 mmol/L | 0.71 (0.22–2.32) | 0.574 | 101 | 42 |
| Composite: achieved K⁺ — index year 2010–2014 | Achieved K⁺ 4.0–4.5 vs <4.0 mmol/L | 1.18 (0.79–1.77) | 0.412 | 561 | 170 |
|  | Achieved K⁺ 4.5–5.0 vs <4.0 mmol/L | 1.15 (0.71–1.85) | 0.575 | 561 | 170 |
|  | Achieved K⁺ ≥5.0 vs <4.0 mmol/L | 1.04 (0.58–1.86) | 0.893 | 561 | 170 |
| Composite: achieved K⁺ — index year 2015–2019 | Achieved K⁺ 4.0–4.5 vs <4.0 mmol/L | 0.74 (0.54–1.02) | 0.064 | 1033 | 239 |
|  | Achieved K⁺ 4.5–5.0 vs <4.0 mmol/L | 0.74 (0.51–1.06) | 0.102 | 1033 | 239 |
|  | Achieved K⁺ ≥5.0 vs <4.0 mmol/L | 1.10 (0.68–1.80) | 0.694 | 1033 | 239 |
| Composite: achieved K⁺ — index year ≥2020 | Achieved K⁺ 4.0–4.5 vs <4.0 mmol/L | 0.94 (0.67–1.33) | 0.735 | 2474 | 172 |
|  | Achieved K⁺ 4.5–5.0 vs <4.0 mmol/L | 0.34 (0.20–0.57) | <0.001 | 2474 | 172 |
|  | Achieved K⁺ ≥5.0 vs <4.0 mmol/L | 0.54 (0.28–1.04) | 0.064 | 2474 | 172 |
| Composite: achieved K⁺ — index era 2000–2014 | Achieved K⁺ 4.0–4.5 vs <4.0 mmol/L | 0.99 (0.70–1.42) | 0.979 | 662 | 212 |
|  | Achieved K⁺ 4.5–5.0 vs <4.0 mmol/L | 0.98 (0.64–1.49) | 0.920 | 662 | 212 |
|  | Achieved K⁺ ≥5.0 vs <4.0 mmol/L | 0.94 (0.56–1.56) | 0.812 | 662 | 212 |
| Composite: achieved K⁺ — index era 2015–2025 | Achieved K⁺ 4.0–4.5 vs <4.0 mmol/L | 0.83 (0.66–1.04) | 0.111 | 3507 | 411 |
|  | Achieved K⁺ 4.5–5.0 vs <4.0 mmol/L | 0.55 (0.41–0.74) | <0.001 | 3507 | 411 |
|  | Achieved K⁺ ≥5.0 vs <4.0 mmol/L | 0.79 (0.53–1.15) | 0.218 | 3507 | 411 |
| Composite: ΔK⁺ — index 2015 onwards | ΔK⁺ tertile 2 vs tertile 1 (lowest) | 0.81 (0.63–1.04) | 0.106 | 3507 | 411 |
|  | ΔK⁺ tertile 3 (highest) vs tertile 1 | 0.66 (0.50–0.88) | 0.004 | 3507 | 411 |

**Supplementary Table S13. Descriptive statistics and interaction tests for calendar period**

| **Item** | **Value** | **Detail** |
| --- | --- | --- |
| index year range | 2000 to 2025 |  |
| P for interaction, achieved K x calendar period | 0.0008057 |  |
| period ≤2009 | n=101, events=42 | median follow-up 10.8 yr |
| period 2010–2014 | n=561, events=170 | median follow-up 10.52 yr |
| period 2015–2019 | n=1033, events=239 | median follow-up 6.07 yr |
| period ≥2020 | n=2474, events=172 | median follow-up 1.78 yr |
| interaction P with ≤2009 dropped | 0.0003815 |  |
| interaction P with 2010–2014 dropped | 0.005262 |  |
| interaction P with 2015–2019 dropped | 0.003075 |  |
| interaction P with ≥2020 dropped | 0.1508 |  |
| interaction P, two-era split | 0.1052 |  |
| interaction P, continuous index year | 0.01053 |  |
| interaction P, exposure x follow-up epoch | 0.02333 |  |

Index years range 2000–2025; median follow-up shortens across periods purely by construction (later cohorts have had less time to accrue events), so the period-by-period gradient (interaction P = 0.0008) needs to be read together with Table S4, which partitions follow-up time rather than calendar period and shows the same pattern of a stronger early association; Table S14 shows that the calendar-period interaction survives adjustment for the follow-up-epoch interaction while the reverse does not; shorter follow-up in recent patients is therefore unlikely to be the whole explanation, but the two cannot be separated in these data. The gradient is, however, carried almost entirely by the ≥2020 period: the interaction is P = 0.1052 on a two-era split and P = 0.1508 when the ≥2020 period is dropped.

**Supplementary Table S14.** Calendar period versus follow-up epoch: disentangling test (n = 4,169)

| **Test** | **P** |
| --- | --- |
| period interaction, after epoch interaction already in the model | 0.012 |
| epoch interaction, after period interaction already in the model | 0.653 |

Joint model with achieved K⁺ × calendar-period and achieved K⁺ × follow-up-epoch interactions both included (survival-split at 3 years). Calendar period still explains outcome variation after the follow-up-epoch interaction is already in the model (P = 0.012); the follow-up-epoch interaction adds nothing detectabke once calendar period is already in the model (P = 0.653) — the opposite of the working hypothesis that the calendar-period gradient (Table S12) was simply an artifact of shorter follow-up in more recently indexed patients. Period carries information epoch does not, consistent with the TriNetX network's own data completeness changing by calendar era

**Supplementary Table S15.** E-value for unmeasured confounding of the primary achieved-potassium association

| **Quantity** | **Value** |
| --- | --- |
| Composite MI/stroke/death — Adjusted hazard ratio | 0.65 (0.52-0.83) |
| Composite MI/stroke/death — Upper confidence limit | 0.83 |
| Composite MI/stroke/death — E-value for the estimate (HR as RR) | 2.43 |
| Composite MI/stroke/death — E-value for the estimate (common-outcome correction) | 2.02 |
| Composite MI/stroke/death — E-value for the confidence limit (common-outcome correction) | 1.54 |
| Incident heart failure — Adjusted hazard ratio | 0.54 (0.36-0.80) |
| Incident heart failure — Upper confidence limit | 0.80 |
| Incident heart failure — E-value for the estimate (HR as RR) | 3.13 |
| Incident heart failure — E-value for the estimate (common-outcome correction) | 2.44 |
| Incident heart failure — E-value for the confidence limit (common-outcome correction) | 1.61 |
| Interpretation  An unmeasured confounder would need associations of at least this magnitude with both the achieved potassium category and the outcome, above the measured covariates, to explain the association away. The common-outcome correction is applied to both outcomes for consistency; the composite outcome is not rare (623/4,169, 14.9%), whereas incident heart failure is less common (248/3,632, 6.8%), for which the correction is conservative. The uncorrected row is an upper bound; the corrected rows apply VanderWeele's approximate hazard-ratio-to-risk-ratio conversion. | |

**Supplementary Table S16.** Achieved K⁺ and ΔK⁺ by whether the patient was biochemically worked up for PA, with the formal interaction test

| **Model** | **Term** | **HR (95% CI)** | **P** |
| --- | --- | --- | --- |
| Composite: achieved K⁺ — worked up for PA | Achieved K⁺ 4.0–4.5 vs <4.0 mmol/L | 0.89 (0.69–1.14) | 0.349 |
|  | Achieved K⁺ 4.5–5.0 vs <4.0 mmol/L | 0.61 (0.44–0.83) | 0.002 |
|  | Achieved K⁺ ≥5.0 vs <4.0 mmol/L | 0.67 (0.44–1.02) | 0.061 |
| Composite: ΔK⁺ — worked up for PA | ΔK⁺ tertile 2 vs tertile 1 (lowest) | 0.83 (0.64–1.09) | 0.183 |
|  | ΔK⁺ tertile 3 (highest) vs tertile 1 | 0.72 (0.53–0.96) | 0.028 |
| Composite: achieved K⁺ — not worked up for PA | Achieved K⁺ 4.0–4.5 vs <4.0 mmol/L | 0.90 (0.67–1.20) | 0.462 |
|  | Achieved K⁺ 4.5–5.0 vs <4.0 mmol/L | 0.73 (0.51–1.06) | 0.097 |
|  | Achieved K⁺ ≥5.0 vs <4.0 mmol/L | 1.01 (0.65–1.58) | 0.959 |
| Composite: ΔK⁺ — not worked up for PA | ΔK⁺ tertile 2 vs tertile 1 (lowest) | 0.77 (0.56–1.06) | 0.106 |
|  | ΔK⁺ tertile 3 (highest) vs tertile 1 | 0.78 (0.55–1.10) | 0.153 |
| Interaction test | Achieved K⁺ × worked-up-for-PA status |  | 0.517 |
|  | ΔK⁺ × worked-up-for-PA status |  | 0.630 |

P for interaction, achieved K⁺ × worked-up status = 0.517; ΔK⁺ × worked-up status = 0.630 (the interaction-test rows carry only a P value, no HR). The association did not differ significantly by whether the patient had a renin or aldosterone measurement on record, although the confidence intervals around the interaction terms do not exclude moderate heterogeneity (ratio of hazard ratios for the 4.5–5.0 category, 0.81, 95% CI 0.50–1.29); PA misclassification in the never-worked-up group is retained as a limitation.

**Supplementary Table S17.** Achieved K⁺ ≥5.0 mmol/L, by baseline eGFR, with the formal interaction test

| **Model** | **Term** | **HR (95% CI)** | **P** |
| --- | --- | --- | --- |
| Composite: achieved K⁺ — baseline eGFR ≥60 | Achieved K⁺ 4.0–4.5 vs <4.0 mmol/L | 1.01 (0.78–1.30) | 0.949 |
|  | Achieved K⁺ 4.5–5.0 vs <4.0 mmol/L | 0.82 (0.60–1.13) | 0.236 |
|  | Achieved K⁺ ≥5.0 vs <4.0 mmol/L | 1.02 (0.66–1.59) | 0.915 |
| Composite: achieved K⁺ — baseline eGFR <60 | Achieved K⁺ 4.0–4.5 vs <4.0 mmol/L | 0.71 (0.53–0.96) | 0.026 |
|  | Achieved K⁺ 4.5–5.0 vs <4.0 mmol/L | 0.49 (0.34–0.70) | <0.001 |
|  | Achieved K⁺ ≥5.0 vs <4.0 mmol/L | 0.61 (0.40–0.94) | 0.024 |
| Interaction test | Achieved K⁺ × baseline eGFR ≥60 |  | 0.216 |

P for interaction = 0.216. The ≥5.0 mmol/L point estimate is not more attenuated in the eGFR<60 subgroup than in eGFR≥60 (if anything the reverse), which does not support renal-impairment channelling as the explanation for the attenuated ≥5.0 category in Table 2 — see also Figure S3.

**Supplementary Table S18.** Achieved K⁺ and the composite outcome, by MRA agent, with the formal interaction test

| **Model** | **Term** | **HR (95% CI)** | **P** |
| --- | --- | --- | --- |
| Composite: achieved K⁺ — spironolactone | Achieved K⁺ 4.0–4.5 vs <4.0 mmol/L | 0.86 (0.70–1.05) | 0.150 |
|  | Achieved K⁺ 4.5–5.0 vs <4.0 mmol/L | 0.61 (0.48–0.79) | <0.001 |
|  | Achieved K⁺ ≥5.0 vs <4.0 mmol/L | 0.79 (0.58–1.08) | 0.143 |
| Composite: achieved K⁺ — eplerenone | Achieved K⁺ 4.0–4.5 vs <4.0 mmol/L | 1.17 (0.62–2.23) | 0.625 |
|  | Achieved K⁺ 4.5–5.0 vs <4.0 mmol/L | 1.07 (0.49–2.36) | 0.866 |
|  | Achieved K⁺ ≥5.0 vs <4.0 mmol/L | 1.45 (0.44–4.77) | 0.545 |
| Interaction test | Achieved K⁺ × MRA agent |  | 0.481 |

P for interaction = 0.481. Eplerenone is only 12% of the cohort (516 patients, 64 events), so its own subgroup estimates are wide — this is a non-significant interaction, not proof the association is identical between agents.

**Supplementary Table S19.** Covariate-standardised absolute risk and risk difference, 3 years from the landmark

| **Outcome** | **Achieved potassium** | **N** | **3-year risk % (95% CI)** | **Standardised risk difference vs <4.0 mmol/L, pp (95% CI)** |
| --- | --- | --- | --- | --- |
| Composite MI/stroke/death | <4.0 | 1360 | 11.31 (9.92–13.15) | reference |
|  | 4.0–4.5 | 1612 | 10.13 (8.84–11.48) | 1.18 (-0.56 to 3.19) |
|  | 4.5–5.0 | 925 | 7.78 (6.43–9.25) | 3.53 (1.71 to 5.77) |
|  | ≥5.0 | 272 | 9.37 (7.51–11.79) | 1.94 (-0.65 to 4.59) |
| Incident heart failure (Fine-Gray) | <4.0 | 1189 | 4.44 (3.43–5.76) | reference |
|  | 4.0–4.5 | 1431 | 3.82 (2.96–4.88) | 0.62 (-0.56 to 1.82) |
|  | 4.5–5.0 | 790 | 2.61 (1.81–3.55) | 1.84 (0.54 to 3.23) |
|  | ≥5.0 | 222 | 3.91 (2.38–5.71) | 0.53 (-1.42 to 2.41) |

Covariate-standardised cumulative incidence, obtained by g-formula standardisation over the adjusted primary model (each patient's risk predicted under every achieved-potassium category and averaged over the observed covariate distribution), with 95% confidence intervals from 500 bootstrap replicates. This replaces the crude Kaplan–Meier / Aalen–Johansen risks reported previously. Standardisation separates the crude and adjusted pictures for the highest category: achieved K⁺ ≥5.0 has a crude 3-year risk of 11.31% against 11.55% in the reference group, while its standardised risk is 9.37% (7.51–11.79) against 11.31%, because that group is older and has worse baseline renal function. The minimum in both outcomes falls in the 4.5–<5.0 mmol/L category. Risk differences are the risk in the <4.0 mmol/L reference group minus the risk in the category shown, so a positive value denotes lower risk in that category.

**Supplementary Table S20. Timing of the exposure-defining potassium measurement, and the association by number of in-window measurements**

| **Panel** | **Item** | **Value** | **HR (95% CI)** | **P** |
| --- | --- | --- | --- | --- |
| A. Timing (day from index) | exposure-defining measurement day | median 121 (IQR 75–157; 10th–90th 35–172) |  |  |
|  | all in-window measurements, day | median 75 (IQR 38–123; 10th–90th 21–157) |  |  |
| B. Association by number of in-window measurements | 1 measurement(s) in window | Achieved K⁺ 4.0–4.5 vs <4.0 mmol/L | 0.80 (0.54–1.18) | 0.254 |
|  |  | Achieved K⁺ 4.5–5.0 vs <4.0 mmol/L | 0.74 (0.46–1.21) | 0.233 |
|  |  | Achieved K⁺ ≥5.0 vs <4.0 mmol/L | 0.76 (0.42–1.38) | 0.364 |
|  | 2 measurement(s) in window | Achieved K⁺ 4.0–4.5 vs <4.0 mmol/L | 0.89 (0.57–1.38) | 0.595 |
|  |  | Achieved K⁺ 4.5–5.0 vs <4.0 mmol/L | 0.50 (0.28–0.90) | 0.021 |
|  |  | Achieved K⁺ ≥5.0 vs <4.0 mmol/L | 1.09 (0.56–2.11) | 0.804 |
|  | 3-4 measurement(s) in window | Achieved K⁺ 4.0–4.5 vs <4.0 mmol/L | 0.77 (0.47–1.26) | 0.304 |
|  |  | Achieved K⁺ 4.5–5.0 vs <4.0 mmol/L | 0.88 (0.52–1.49) | 0.642 |
|  |  | Achieved K⁺ ≥5.0 vs <4.0 mmol/L | 0.46 (0.20–1.07) | 0.071 |
|  | >=5 measurement(s) in window | Achieved K⁺ 4.0–4.5 vs <4.0 mmol/L | 1.05 (0.77–1.45) | 0.740 |
|  |  | Achieved K⁺ 4.5–5.0 vs <4.0 mmol/L | 0.55 (0.36–0.86) | 0.008 |
|  |  | Achieved K⁺ ≥5.0 vs <4.0 mmol/L | 0.89 (0.53–1.50) | 0.668 |

Panel A contrasts the date of the measurement that defines the exposure (the last value in days 14–180) with the date of a typical in-window measurement; the two differ substantially. Panel B repeats the primary model within strata of the number of in-window measurements. P for interaction, achieved K⁺ × (≥3 in-window measurements) = 0.64.

**Supplementary Table S21. Tests for a non-linear or J-shaped association with achieved potassium**

| **Test** | **Specification** | **P** |
| --- | --- | --- |
| linear achieved K+ vs no exposure |  | 0.004 |
| best FP1 vs linear | power -1 | 0.510 |
| best FP2 vs best FP1 | powers -2, 3 | 0.977 |
| extra-knot spline (incl. knot at 5.0) vs linear | knots 4, 4.4, 5 | 0.235 |
| restricted cubic spline (3 df, default knots; Figure S3 model) vs linear |  | 0.622 |

Interior knots were placed at the tertiles of achieved potassium (4.0 and 4.4 mmol/L), with a third at 5.0 mmol/L in the extra-knot model. A natural spline is constrained to be linear only beyond its boundary knots. These are global tests of fit and indicate whether a linear term fits worse, not where any departure lies. Neither improves on a linear term for the composite endpoint. The same tests fitted to incident heart failure give a different answer; see Supplementary Table S26. The restricted cubic spline row above is the model fitted for Supplementary Figure S3 (natural spline, 3 degrees of freedom, default knot placement); it is a different specification from the fractional-polynomial and extra-knot rows above it. Its own 'any association' test (spline vs no exposure, P = 0.024, 3 degrees of freedom, given in the Figure S3 legend) is therefore not the same test as the 'linear achieved K+ vs no exposure' row above (P = 0.004, 1 degree of freedom): the two P values test different hypotheses about the same relationship, not inconsistent estimates of the same one.

**Supplementary Table S22. Selection into the analytic cohort: standardised differences and inverse-probability-of-selection weighting**

| **Panel** | **Item** | **Value** | **HR (95% CI)** | **P** | **SMD** |
| --- | --- | --- | --- | --- | --- |
| A. Standardised differences, analytic vs excluded | age | 59.028 vs 59.394 |  |  | -0.026 |
|  | k0_last | 3.799 vs 3.789 |  |  | 0.017 |
|  | egfr0 | 75.971 vs 67.200 |  |  | 0.306 |
|  | htn | 0.868 vs 0.921 |  |  | -0.174 |
|  | dm | 0.307 vs 0.414 |  |  | -0.225 |
|  | ckd | 0.253 vs 0.450 |  |  | -0.421 |
|  | hf | 0.123 vs 0.266 |  |  | -0.368 |
|  | af | 0.095 vs 0.145 |  |  | -0.155 |
|  | ihd | 0.197 vs 0.349 |  |  | -0.347 |
|  | cvd | 0.127 vs 0.279 |  |  | -0.384 |
|  | obes | 0.339 vs 0.391 |  |  | -0.107 |
| B. Composite outcome, achieved potassium | unweighted (same rows) | Achieved K⁺ 4.0–4.5 vs <4.0 mmol/L | 0.88 (0.73–1.07) | 0.191 |  |
|  |  | Achieved K⁺ 4.5–5.0 vs <4.0 mmol/L | 0.65 (0.52–0.83) | <0.001 |  |
|  |  | Achieved K⁺ ≥5.0 vs <4.0 mmol/L | 0.81 (0.59–1.09) | 0.159 |  |
|  | IPW for selection | Achieved K⁺ 4.0–4.5 vs <4.0 mmol/L | 0.89 (0.73–1.08) | 0.244 |  |
|  |  | Achieved K⁺ 4.5–5.0 vs <4.0 mmol/L | 0.64 (0.50–0.81) | <0.001 |  |
|  |  | Achieved K⁺ ≥5.0 vs <4.0 mmol/L | 0.79 (0.58–1.07) | 0.128 |  |

Weights are the inverse of the fitted probability of inclusion, estimated among all 5,463 patients who reached the paired-potassium step (4,169 included, 1,294 excluded) and trimmed at the 99th centile (range 1.08–1.88, mean 1.21). The excluded group is substantially sicker, but weighting for that selection does not materially change the estimate.

**Supplementary Table S23. Negative control outcomes ascertained on the primary outcome's pathway**

| **Negative control** | **Term** | **HR (95% CI)** | **P** |
| --- | --- | --- | --- |
| cellulitis | ΔK⁺ tertile 2 vs tertile 1 (lowest) | 0.89 (0.58–1.37) | 0.604 |
|  | ΔK⁺ tertile 3 (highest) vs tertile 1 | 0.59 (0.35–0.99) | 0.046 |
| diverticulitis | ΔK⁺ tertile 2 vs tertile 1 (lowest) | 0.91 (0.60–1.38) | 0.663 |
|  | ΔK⁺ tertile 3 (highest) vs tertile 1 | 0.90 (0.58–1.41) | 0.652 |
| cholecystitis | ΔK⁺ tertile 2 vs tertile 1 (lowest) | 1.14 (0.67–1.94) | 0.619 |
|  | ΔK⁺ tertile 3 (highest) vs tertile 1 | 1.02 (0.56–1.83) | 0.951 |

Unlike cataract, herpes zoster and inguinal hernia, these three outcomes are required to be coded at an inpatient, acute-care or emergency encounter, exactly as the primary outcome is; they therefore probe differential outcome capture across the network rather than general confounding. Adjustment set as for the primary model.

**Supplementary Table S24. Change in potassium and the composite outcome, stratified by MRA fill count**

| **Stratum** | **Term** | **HR (95% CI)** | **P** |
| --- | --- | --- | --- |
| MRA fills 1 | ΔK⁺ tertile 2 vs tertile 1 (lowest) | 1.05 (0.66–1.65) | 0.847 |
|  | ΔK⁺ tertile 3 (highest) vs tertile 1 | 1.13 (0.63–2.01) | 0.687 |
| MRA fills 2-3 | ΔK⁺ tertile 2 vs tertile 1 (lowest) | 0.83 (0.57–1.22) | 0.353 |
|  | ΔK⁺ tertile 3 (highest) vs tertile 1 | 0.74 (0.48–1.13) | 0.162 |
| MRA fills 4-6 | ΔK⁺ tertile 2 vs tertile 1 (lowest) | 0.40 (0.24–0.69) | <0.001 |
|  | ΔK⁺ tertile 3 (highest) vs tertile 1 | 0.33 (0.19–0.56) | <0.001 |
| MRA fills >=7 | ΔK⁺ tertile 2 vs tertile 1 (lowest) | 0.71 (0.42–1.18) | 0.186 |
|  | ΔK⁺ tertile 3 (highest) vs tertile 1 | 0.75 (0.43–1.29) | 0.297 |

Fill count over the first 180 days is measured in the same window as the exposure, so conditioning on it in any form -- by adjustment or by stratification -- conditions on a post-baseline variable; stratification makes that conditioning explicit rather than avoiding it. The estimates differ across strata without a monotone trend and the formal interaction test is not significant (P = 0.276). The four strata contain 3,267 of the 4,169 patients: fill counts come from a dispensing table that is joined to the cohort, and 902 patients have no matching record. Supplementary Table S6, which adjusts for fill count rather than stratifying on it, retains all 4,169 by coding those patients as zero fills and pooling them with the single-fill reference; the two tables therefore treat the same 902 patients differently. P for interaction, ΔK⁺ × fill-count stratum = 0.276.

**Supplementary Table S25. Proportional-hazards test for the incident-heart-failure model**

| **Model** | **Term** | **chisq** | **df** | **P** |
| --- | --- | --- | --- | --- |
| Incident heart failure | Achieved potassium category | 2.145 | 3 | 0.5429 |
|  | Age, per year | 0.736 | 1 | 0.3910 |
|  | Sex | 0.448 | 1 | 0.5032 |
|  | race_f | 2.826 | 3 | 0.4192 |
|  | MRA agent (spironolactone vs eplerenone) | 1.207 | 1 | 0.2719 |
|  | Baseline potassium, per mmol/L | 0.585 | 1 | 0.4445 |
|  | Baseline eGFR, per mL/min/1.73m² | 1.557 | 1 | 0.2122 |
|  | Hypertension | 0.03 | 1 | 0.8627 |
|  | Diabetes | 0.009 | 1 | 0.9243 |
|  | Chronic kidney disease | 0.592 | 1 | 0.4415 |
|  | Atrial fibrillation | 0.144 | 1 | 0.7046 |
|  | Ischaemic heart disease | 0.571 | 1 | 0.4498 |
|  | ACE inhibitor | 1.805 | 1 | 0.1791 |
|  | ARB | 4.569 | 1 | 0.03255 |
|  | Thiazide | 0.021 | 1 | 0.8847 |
|  | Loop diuretic | 0.08 | 1 | 0.7772 |
|  | Potassium supplement | 1.768 | 1 | 0.1836 |
| Incident heart failure | Global test | 20.71 | 21 | 0.4768 |

Scaled Schoenfeld residual test for every term in the adjusted cause-specific model (n = 3,632 with 248 events). The exposure term gives P = 0.543 and the global test P = 0.477, so there is no evidence against proportional hazards for the exposure or for the model as a whole. This is not proof that each hazard ratio is constant: the angiotensin receptor blocker term gives P = 0.033, and a non-significant test is not evidence of no violation.

**Supplementary Table S26. Tests for a non-linear association with achieved potassium and incident heart failure**

| **Comparison** | **chisq** | **df** | **P** |
| --- | --- | --- | --- |
| Any association with achieved potassium (linear vs none) | 7.005 | 1 | 0.008127 |
| Restricted cubic spline vs linear (knots at the 10th, 50th, 90th centiles) | 7.032 | 3 | 0.07089 |
| Spline with an added knot at 5.0 mmol/L vs linear | 13.055 | 4 | 0.01101 |
| Best one-term fractional polynomial (power 3) vs linear | 0.793 | 1 | 0.3732 |
| Best two-term fractional polynomial (powers -2, -2) vs one-term | 3.059 | 2 | 0.2166 |

The same comparison as Supplementary Table S21, fitted to incident heart failure rather than to the composite endpoint. The spline comparisons are nested likelihood-ratio tests. The fractional-polynomial rows are not: the best one- and two-term powers are chosen by search over the conventional set (-2, -1, -0.5, 0, 0.5, 1, 2, 3) with a maximum degree of two, and the deviance differences are referred to one and two degrees of freedom as in the function-selection procedure. A restricted cubic spline does not improve on a linear term (P = 0.071), but a spline carrying an additional interior knot at 5.0 mmol/L does (P = 0.011); the fractional-polynomial comparisons do not improve on a linear term either (best one-term polynomial versus linear, P = 0.373; best two-term versus one-term, P = 0.217). Only the spline given an explicit knot at 5.0 mmol/L improves on a linear term, and all of these are global tests of fit that do not locate where any departure lies. The first row tests whether there is any association with achieved potassium, not whether it is non-linear. The spline used interior knots at the 10th, 50th and 90th centiles of achieved potassium, with a fourth added at 5.0 mmol/L; a natural spline is constrained to be linear only beyond its boundary knots. This does not amount to a reversal. The categorical contrast the question turns on, ≥5.0 versus 4.5–<5.0 mmol/L read off the four-category model that produces the primary estimates, is 1.45 (0.86 to 2.46), P = 0.166 on 23 events in the highest category. A J-shaped reversal is suggested by this point estimate and by the one spline specification given a knot at 5.0 mmol/L, and is not established.

**Supplementary Table S27. Composition of the cohort by hyperaldosteronism diagnosis code**

| **Group** | **n** | **%** |
| --- | --- | --- |
| Analytic cohort | 4169 | 100.0 |
| Any E26 record in the extracted diagnosis table | 3872 | 92.9 |
| At least one specific primary-aldosteronism code (E26.0, .01, .02, .09) | 2068 | 49.6 |
| No specific primary code; unspecified hyperaldosteronism (E26.9) only | 1804 | 43.3 |
| Secondary hyperaldosteronism (E26.1) and no specific primary code | 0 | 0.0 |
| Bartter syndrome (E26.81) and no specific primary code | 0 | 0.0 |
| No E26 record in the extracted diagnosis table | 297 | 7.1 |

Patients carrying E26.1 (secondary hyperaldosteronism) or E26.81 (Bartter syndrome) are excluded at cohort construction and therefore appear as zero. Half the remaining cohort carries only an unspecified code, which is the principal limitation of an administrative case definition. Coding granularity is a property of the documenting clinician and of how complete a patient's record is in this network — 297 patients have no E26 row at all — not a property of the patient, so it is described here rather than used as a stratifying variable.

**Supplementary Table S28. Primary models under a stricter diagnosis requirement**

| **Outcome** | **Cohort** | **Achieved potassium vs <4.0 mmol/L** | **Adjusted HR (95% CI)** |
| --- | --- | --- | --- |
| Composite MI/stroke/death | At least two recorded PA diagnoses | 4.0–<4.5 mmol/L | 0.87 (0.68 to 1.09) |
|  |  | 4.5–<5.0 mmol/L | 0.58 (0.43 to 0.78) |
|  |  | ≥5.0 mmol/L | 0.65 (0.45 to 0.95) |
|  | Primary analysis | 4.0–<4.5 mmol/L | 0.88 (0.73 to 1.07) |
|  |  | 4.5–<5.0 mmol/L | 0.65 (0.52 to 0.83) |
|  |  | ≥5.0 mmol/L | 0.80 (0.59 to 1.09) |
| Incident heart failure | At least two recorded PA diagnoses | 4.0–<4.5 mmol/L | 0.81 (0.58 to 1.14) |
|  |  | 4.5–<5.0 mmol/L | 0.56 (0.36 to 0.88) |
|  |  | ≥5.0 mmol/L | 0.76 (0.44 to 1.30) |
|  | Primary analysis | 4.0–<4.5 mmol/L | 0.83 (0.62 to 1.12) |
|  |  | 4.5–<5.0 mmol/L | 0.54 (0.36 to 0.80) |
|  |  | ≥5.0 mmol/L | 0.78 (0.48 to 1.27) |

Requiring two or more separately dated PA diagnoses probes diagnostic certainty rather than coding granularity: a single stray code is more likely to be provisional or erroneous than a diagnosis carried across two encounters. It retains 3,360 of 4,169 patients (80.6%), and neither estimate moves materially despite the loss of a third of the events.

**Supplementary Table S29. The reference category split, the normal range alone, and contrasts between adjacent categories**

| **Analysis** | **Term** | **HR** | **P** |
| --- | --- | --- | --- |
| A. Composite, reference 3.5-<4.0 | <3.5 | 1.09 (0.81 to 1.47) | 0.580 |
|  | 4.0–4.5 | 0.90 (0.73 to 1.11) | 0.329 |
|  | 4.5–5.0 | 0.67 (0.52 to 0.86) | 0.002 |
|  | ≥5.0 | 0.82 (0.60 to 1.13) | 0.225 |
| A. Incident HF, reference 3.5-<4.0 | <3.5 | 1.04 (0.65 to 1.67) | 0.854 |
|  | 4.0–4.5 | 0.84 (0.61 to 1.16) | 0.297 |
|  | 4.5–5.0 | 0.54 (0.36 to 0.83) | 0.004 |
|  | ≥5.0 | 0.79 (0.48 to 1.31) | 0.356 |
| B. Composite, normal range only (3.5-<5.0) | 4.0–4.5 | 0.91 (0.74 to 1.12) | 0.369 |
|  | 4.5–5.0 | 0.68 (0.53 to 0.88) | 0.003 |
| B. Incident HF, normal range only (3.5-<5.0) | 4.0–4.5 | 0.86 (0.62 to 1.19) | 0.367 |
|  | 4.5–5.0 | 0.54 (0.36 to 0.83) | 0.005 |
| C. Composite, reference 4.0_4.5 | <4.0 | 1.14 (0.94 to 1.38) | 0.191 |
|  | 4.5–5.0 | 0.74 (0.59 to 0.93) | 0.010 |
|  | ≥5.0 | 0.91 (0.68 to 1.23) | 0.550 |
| C. Incident HF, reference 4.0_4.5 | <4.0 | 1.20 (0.89 to 1.62) | 0.221 |
|  | 4.5–5.0 | 0.65 (0.44 to 0.95) | 0.025 |
|  | ≥5.0 | 0.94 (0.58 to 1.51) | 0.794 |
| C. Composite, reference 4.5_5.0 | <4.0 | 1.53 (1.21 to 1.94) | <0.001 |
|  | 4.0–4.5 | 1.35 (1.07 to 1.69) | 0.010 |
|  | ≥5.0 | 1.23 (0.90 to 1.69) | 0.200 |
| C. Incident HF, reference 4.5_5.0 | <4.0 | 1.86 (1.25 to 2.78) | 0.002 |
|  | 4.0–4.5 | 1.55 (1.06 to 2.27) | 0.025 |
|  | ≥5.0 | 1.45 (0.86 to 2.46) | 0.166 |

The reference category of <4.0 mmol/L pools frank hypokalaemia with low-normal potassium. Panel A splits it at 3.5 mmol/L: relative to 3.5–<4.0 mmol/L, achieved potassium below 3.5 mmol/L gave 1.09 (0.81 to 1.47) for the composite outcome and 1.04 (0.65 to 1.67) for heart failure. These intervals are wide and do not exclude a difference between the two halves of the reference category; what they do show is that the gradient is not produced by frank hypokalaemia alone, since it survives in Panel B. Panel B repeats the analysis within 3.5 to <5.0 mmol/L, where the gradient persists. Panel C gives the contrasts between adjacent categories: 4.5–<5.0 versus 4.0–<4.5 mmol/L is 0.74 (0.59 to 0.93) for the composite outcome and 0.65 (0.44 to 0.95) for incident heart failure, so the two adjacent normal-range categories do differ. Taking 4.5–<5.0 mmol/L as the reference instead, the ≥5.0 mmol/L contrast is 1.23 (0.90 to 1.69) and 1.45 (0.86 to 2.46), neither significant: the lowest category is separated from those below it but not from the one above it.

**Supplementary Table S30. Sequential adjustment**

| **Outcome** | **Step** | **4.0–<4.5** | **4.5–<5.0** | **≥5.0** | **P for ≥5.0** |
| --- | --- | --- | --- | --- | --- |
| Composite MI/stroke/death | 1 crude | 0.93 (0.77 to 1.12) | 0.81 (0.64 to 1.01) | 1.21 (0.91 to 1.62) | 0.190 |
|  | 2 + age, sex, race | 0.85 (0.71 to 1.03) | 0.68 (0.54 to 0.86) | 0.96 (0.72 to 1.29) | 0.796 |
|  | 3 + baseline K+ | 0.84 (0.70 to 1.02) | 0.67 (0.53 to 0.84) | 0.94 (0.69 to 1.26) | 0.662 |
|  | 4 + baseline eGFR, CKD | 0.84 (0.70 to 1.02) | 0.65 (0.51 to 0.82) | 0.79 (0.58 to 1.06) | 0.116 |
|  | 5 + prevalent disease (strata) | 0.86 (0.71 to 1.04) | 0.64 (0.50 to 0.81) | 0.80 (0.59 to 1.08) | 0.146 |
|  | 6 + remaining comorbidity | 0.86 (0.71 to 1.04) | 0.66 (0.52 to 0.83) | 0.80 (0.59 to 1.08) | 0.143 |
|  | 7 + agent, co-medications (full) | 0.88 (0.73 to 1.07) | 0.65 (0.52 to 0.83) | 0.80 (0.59 to 1.09) | 0.159 |
| Incident heart failure | 1 crude | 0.93 (0.70 to 1.24) | 0.64 (0.44 to 0.93) | 1.24 (0.79 to 1.97) | 0.350 |
|  | 2 + age, sex, race | 0.88 (0.66 to 1.17) | 0.55 (0.38 to 0.81) | 1.00 (0.63 to 1.59) | 0.995 |
|  | 3 + baseline K+ | 0.88 (0.65 to 1.18) | 0.55 (0.37 to 0.82) | 1.00 (0.62 to 1.61) | 0.995 |
|  | 4 + baseline eGFR, CKD | 0.85 (0.63 to 1.14) | 0.53 (0.35 to 0.78) | 0.78 (0.49 to 1.27) | 0.320 |
|  | 5 + prevalent disease (strata) | 0.86 (0.64 to 1.15) | 0.54 (0.36 to 0.80) | 0.72 (0.44 to 1.18) | 0.195 |
|  | 6 + remaining comorbidity | 0.85 (0.63 to 1.13) | 0.56 (0.37 to 0.83) | 0.77 (0.47 to 1.26) | 0.289 |
|  | 7 + agent, co-medications (full) | 0.83 (0.62 to 1.12) | 0.54 (0.36 to 0.80) | 0.78 (0.48 to 1.27) | 0.321 |

Covariates enter in the order shown. Step 1 is the crude model reported in Table 2, with no covariates and no strata, so the chain begins at the same value the main table carries. The strata enter at step 5. Race enters with the other demographics at step 2.

**Supplementary Table S31. Race by achieved potassium, and the primary models estimated with and without race**

| **Panel** | **Item** | **<4.0** | **4.0–<4.5** | **4.5–<5.0** | **≥5.0** | **Overall** |
| --- | --- | --- | --- | --- | --- | --- |
| A. Distribution by achieved potassium | White | 532 (39.1) | 756 (46.9) | 482 (52.1) | 140 (51.5) | 1910 (45.8) |
|  | Asian | 97 (7.1) | 100 (6.2) | 62 (6.7) | 18 (6.6) | 277 (6.6) |
|  | Black or African American | 570 (41.9) | 537 (33.3) | 249 (26.9) | 73 (26.8) | 1429 (34.3) |
|  | Other or Unknown | 161 (11.8) | 219 (13.6) | 132 (14.3) | 41 (15.1) | 553 (13.3) |
| B. Adjusted HR for achieved potassium, with and without race |  | with race | without race |  |  |  |
|  | composite — Achieved K⁺ 4.0–4.5 vs <4.0 mmol/L | 0.88 | 0.873 |  |  |  |
|  | composite — Achieved K⁺ 4.5–5.0 vs <4.0 mmol/L | 0.654 | 0.64 |  |  |  |
|  | composite — Achieved K⁺ ≥5.0 vs <4.0 mmol/L | 0.805 | 0.796 |  |  |  |
|  | heart failure — Achieved K⁺ 4.0–4.5 vs <4.0 mmol/L | 0.831 | 0.847 |  |  |  |
|  | heart failure — Achieved K⁺ 4.5–5.0 vs <4.0 mmol/L | 0.537 | 0.532 |  |  |  |
|  | heart failure — Achieved K⁺ ≥5.0 vs <4.0 mmol/L | 0.78 | 0.747 |  |  |  |
|  | Likelihood-ratio test for race, composite outcome | P = 0.051 |  |  |  |  |

**Supplementary Table S32. Outcome ascertainment: acute-care encounters only versus any encounter type**

| **Model** | **Outcome** | **Term** | **HR (95% CI)** |
| --- | --- | --- | --- |
| capture by exposure category | patients with >=1 outcome code | Achieved K⁺ <4.0 | 232/415 (55.9% with an acute-care code) |
|  |  | Achieved K⁺ 4.0–4.5 | 250/454 (55.1% with an acute-care code) |
|  |  | Achieved K⁺ 4.5–5.0 | 133/271 (49.1% with an acute-care code) |
|  |  | Achieved K⁺ ≥5.0 | 63/101 (62.4% with an acute-care code) |
| acute-care only (primary) | composite | Achieved K⁺ 4.0–4.5 | 0.88 (0.73–1.07) |
|  |  | Achieved K⁺ 4.5–5.0 | 0.65 (0.52–0.83) |
|  |  | Achieved K⁺ ≥5.0 | 0.81 (0.59–1.09) |
| any encounter | composite | Achieved K⁺ 4.0–4.5 | 0.78 (0.65–0.94) |
|  |  | Achieved K⁺ 4.5–5.0 | 0.66 (0.53–0.83) |
|  |  | Achieved K⁺ ≥5.0 | 0.79 (0.59–1.06) |
| acute-care only (primary) | heart failure | Achieved K⁺ 4.0–4.5 | 0.83 (0.62–1.12) |
|  |  | Achieved K⁺ 4.5–5.0 | 0.54 (0.36–0.80) |
|  |  | Achieved K⁺ ≥5.0 | 0.78 (0.48–1.27) |
| any encounter | heart failure | Achieved K⁺ 4.0–4.5 | 0.92 (0.71–1.17) |
|  |  | Achieved K⁺ 4.5–5.0 | 0.75 (0.55–1.02) |
|  |  | Achieved K⁺ ≥5.0 | 0.95 (0.64–1.42) |
| any encounter | event counts | composite 623 → 686; incident heart failure 248 → 373 |  |

**Supplementary Table S33. Primary models restricted to patients carrying a diagnosis code specific to primary aldosteronism**

| **Cohort** | **Outcome** | **4.0–<4.5** | **4.5–<5.0** | **≥5.0** | **n** | **Events** |
| --- | --- | --- | --- | --- | --- | --- |
| Primary analysis (all E26) | composite | 0.88 (0.73 to 1.07) | 0.65 (0.52 to 0.83) | 0.80 (0.59 to 1.09) | 4169 | 623 |
|  | heart failure | 0.83 (0.62 to 1.12) | 0.54 (0.36 to 0.80) | 0.78 (0.48 to 1.27) | 3632 | 248 |
| Specific PA code at any time | composite | 0.93 (0.69 to 1.26) | 0.66 (0.45 to 0.96) | 0.73 (0.45 to 1.18) | 2068 | 258 |
|  | heart failure | 0.70 (0.43 to 1.14) | 0.80 (0.45 to 1.43) | 1.09 (0.56 to 2.11) | 1836 | 107 |
| Specific PA code on or before the landmark | composite | 0.96 (0.67 to 1.36) | 0.66 (0.42 to 1.01) | 0.74 (0.42 to 1.29) | 1784 | 195 |
|  | heart failure | 0.51 (0.26 to 1.00) | 1.02 (0.51 to 2.02) | 1.24 (0.52 to 2.93) | 1567 | 64 |

Adjusted cause-specific hazard ratios (95% CI) for achieved potassium relative to <4.0 mmol/L, from the primary model refitted within each cohort (adjusted for age, sex, race, MRA agent, baseline potassium and eGFR, hypertension, diabetes, chronic kidney disease, atrial fibrillation, ischemic heart disease and concomitant medications; stratified by prior cerebrovascular disease and, for the composite outcome, prior heart failure). Specific PA codes are E26.0, E26.01, E26.02 and E26.09. n and Events refer to each outcome's analytic cohort; heart-failure cohorts exclude patients with heart failure at baseline or before the landmark. In the restricted heart-failure cohorts, events across the four achieved-potassium categories (<4.0, 4.0–<4.5, 4.5–<5.0, ≥5.0 mmol/L) were 35, 35, 22 and 15 for a specific code at any time and 22, 16, 17 and 9 for a specific code on or before the landmark.

**Supplementary Table S34. Sensitivity to adjustment for the concurrent change in eGFR**

| **Outcome** | **Model** | **Cohort** | **ΔeGFR in the model** | **Term** | **HR (95% CI)** | **N** | **Events** |
| --- | --- | --- | --- | --- | --- | --- | --- |
| Composite MI/stroke/death | Achieved potassium | Full cohort | No | Achieved K⁺ 4.0–<4.5 vs <4.0 mmol/L | 0.88 (0.73 to 1.07) | 4169 | 623 |
|  |  |  | No | Achieved K⁺ 4.5–<5.0 vs <4.0 mmol/L | 0.65 (0.52 to 0.83) | 4169 | 623 |
|  |  |  | No | Achieved K⁺ ≥5.0 vs <4.0 mmol/L | 0.80 (0.59 to 1.09) | 4169 | 623 |
|  |  | Subset with a paired creatinine | No | Achieved K⁺ 4.0–<4.5 vs <4.0 mmol/L | 0.89 (0.73 to 1.07) | 4111 | 610 |
|  |  |  | No | Achieved K⁺ 4.5–<5.0 vs <4.0 mmol/L | 0.65 (0.51 to 0.83) | 4111 | 610 |
|  |  |  | No | Achieved K⁺ ≥5.0 vs <4.0 mmol/L | 0.80 (0.59 to 1.09) | 4111 | 610 |
|  |  |  | Yes | Achieved K⁺ 4.0–<4.5 vs <4.0 mmol/L | 0.86 (0.70 to 1.04) | 4111 | 610 |
|  |  |  | Yes | Achieved K⁺ 4.5–<5.0 vs <4.0 mmol/L | 0.61 (0.48 to 0.78) | 4111 | 610 |
|  |  |  | Yes | Achieved K⁺ ≥5.0 vs <4.0 mmol/L | 0.72 (0.52 to 1.00) | 4111 | 610 |
|  | ΔK⁺ tertile | Full cohort | No | ΔK⁺ tertile 2 vs tertile 1 (lowest) | 0.80 (0.66 to 0.98) | 4169 | 623 |
|  |  |  | No | ΔK⁺ tertile 3 (highest) vs tertile 1 | 0.75 (0.60 to 0.94) | 4169 | 623 |
|  |  | Subset with a paired creatinine | No | ΔK⁺ tertile 2 vs tertile 1 (lowest) | 0.79 (0.64 to 0.97) | 4111 | 610 |
|  |  |  | No | ΔK⁺ tertile 3 (highest) vs tertile 1 | 0.75 (0.60 to 0.94) | 4111 | 610 |
|  |  |  | Yes | ΔK⁺ tertile 2 vs tertile 1 (lowest) | 0.77 (0.62 to 0.94) | 4111 | 610 |
|  |  |  | Yes | ΔK⁺ tertile 3 (highest) vs tertile 1 | 0.71 (0.56 to 0.89) | 4111 | 610 |

Adjusted cause-specific hazard ratios (95% CI) for the composite outcome of myocardial infarction, stroke or death, from the primary adjustment set. ΔeGFR is the change in eGFR (CKD-EPI 2021) from the baseline window to the exposure window (days 14–180), entered as a continuous term. The subset with a paired creatinine comprises the 4,111 patients (610 events) with creatinine in both windows; comparing its rows with and without ΔeGFR separates the effect of adding ΔeGFR from the effect of restricting the sample. Estimates with ΔeGFR in the model are multivariable-adjusted associations, not direct effects.

**Supplementary Table S35. The PAMO potassium threshold of 3.6 mmol/L**

| **Analysis** | **Outcome** | **Achieved potassium (mmol/L) or response** | **N** | **Events** | **HR (95% CI)** |
| --- | --- | --- | --- | --- | --- |
| Achieved potassium dichotomised at 3.6 mmol/L (full cohort) | Composite MI/stroke/death | ≥3.6 | 3720 | 541 | 1.00 (reference) |
|  |  | <3.6 | 449 | 82 | 1.22 (0.96 to 1.56) |
|  | Incident heart failure | ≥3.6 | 3246 | 213 | 1.00 (reference) |
|  |  | <3.6 | 386 | 35 | 1.34 (0.91 to 1.96) |
| Achieved potassium in five categories (full cohort) | Composite MI/stroke/death | <3.6 | 449 | 82 | 1.06 (0.80 to 1.41) |
|  |  | 3.6–<4.0 | 911 | 131 | 1.00 (reference) |
|  |  | 4.0–<4.5 | 1612 | 236 | 0.90 (0.72 to 1.12) |
|  |  | 4.5–<5.0 | 925 | 116 | 0.67 (0.52 to 0.86) |
|  |  | ≥5.0 | 272 | 58 | 0.82 (0.60 to 1.13) |
|  | Incident heart failure | <3.6 | 386 | 35 | 1.11 (0.72 to 1.73) |
|  |  | 3.6–<4.0 | 803 | 54 | 1.00 (reference) |
|  |  | 4.0–<4.5 | 1431 | 98 | 0.86 (0.62 to 1.21) |
|  |  | 4.5–<5.0 | 790 | 38 | 0.56 (0.36 to 0.85) |
|  |  | ≥5.0 | 222 | 23 | 0.81 (0.48 to 1.35) |
| PAMO potassium response (baseline potassium <3.6 mmol/L) | Composite MI/stroke/death | Corrected without supplementation | 546 | 60 | 1.00 (reference) |
|  |  | Corrected with supplementation | 562 | 104 | 1.13 (0.80 to 1.60) |
|  |  | Not corrected | 278 | 45 | 1.03 (0.68 to 1.56) |
|  | Incident heart failure | Corrected without supplementation | 501 | 27 | 1.00 (reference) |
|  |  | Corrected with supplementation | 459 | 34 | 1.06 (0.61 to 1.86) |
|  |  | Not corrected | 241 | 22 | 1.54 (0.84 to 2.84) |

Adjusted cause-specific hazard ratios (95% CI). PAMO, Primary Aldosteronism Medical Treatment Outcome consensus, which defines correction of hypokalaemia as a serum potassium of at least 3.6 mmol/L; the Primary Aldosteronism Surgical Outcome (PASO) consensus uses the same threshold. The dichotomised and five-category models use the primary adjustment set. The PAMO potassium response is estimated among the 1,386 patients with a baseline potassium below 3.6 mmol/L (1,201 free of heart failure at baseline and through the landmark): corrected is an achieved potassium of at least 3.6 mmol/L, and supplementation is a potassium chloride, citrate or bicarbonate prescription between the index date and the landmark; in this analysis a potassium-supplement prescription in the 180 days before the index date replaces the primary supplement covariate. Heart-failure models are restricted to patients free of heart failure at baseline and through the landmark and are not stratified on prevalent heart failure. MI, myocardial infarction.
